# Episodic memory retrieval with increasing task demand: Associations with age, *APOE4* genotype, and Alzheimer’s disease pathology

**DOI:** 10.64898/2026.01.30.26344953

**Authors:** Finja Askevold, Beate Schumann-Werner, Niklas Behrenbruch, Svenja Schwarck, Eóin N. Molloy, Jonathan E. Peelle, Ryan M. O’Leary, Arthur Wingfield, Gusalija Behnisch, Constanze Seidenbecher, Björn H. Schott, Bárbara Morgado, Hermann Esselmann, Jens Wiltfang, Emrah Düzel, Anne Maass, Larissa Fischer

## Abstract

**BACKGROUND:** Episodic memory declines early with aging, reflecting reduced neural resources and diminished memory specificity. However, few studies have created a cognitive challenge with multiple levels of task demand to investigate this early subtle decline. Furthermore, it is unclear whether the genetic Alzheimer’s disease risk factor *APOE4* and early Alzheimer’s pathology constrain the neural resources required to cope with increasing task demands.

**METHODS:** In this preregistered behavioral study named EMCOMP (Episodic Memory & COMPensation), we conducted a semantic episodic memory retrieval task using a sentence-based task demand manipulation with five demand levels at recognition (novel foils, “old” target sentences, and three lure levels) and two demand levels at free and cued recall (gist and details). We collected data from 100 cognitively unimpaired adults (37 young (mean age 24 years), 63 older (mean age 72 years)) with additional neuropsychological testing and blood-based measures available for the older group. First, we investigated differences in EMCOMP episodic memory retrieval accuracy and confidence between young and older adults in linear mixed-effects models. Second, we investigated differences associated with the *APOE4* genotype and plasma-derived Alzheimer’s pathology. Third, we assessed correlations between EMCOMP recognition and recall and established cognitive tests.

**RESULTS:** Young adults showed higher recognition accuracy and confidence as well as a higher recall score and a lower recall error rate compared to older adults. As recognition-task demand increased there was a steeper decline in recognition accuracy and a steeper increase in high-confidence errors in older compared to younger adults and in older *APOE4* carriers compared to older non-carriers. However, we found no associations with Alzheimer’s pathology. EMCOMP performance was positively correlated with established cognitive tests.

**CONCLUSION:** Our study demonstrates age– and *APOE4*-related differences in episodic memory retrieval under increasing task demands – differences that may not be detectable in paradigms with only one or two levels of task demand. By manipulating semantic retrieval specificity, we provide a novel approach to detect subtle cognitive deficits and potential functional compensation in cognitively unimpaired older adults with risk factors or early Alzheimer’s pathology. Future research should extend this work to more diverse samples and combine behavioral assessment with fMRI to examine underlying brain activation patterns and functional connectivity.

## 1. Introduction

Cognitive decline occurs with aging, especially when Alzheimer’s disease risk factors or early Alzheimer’s pathology are present^1,2^. Episodic memory is among the first cognitive functions to decline^3,4^, accompanied by a decline in working memory capacity^5,6^, whereas language processing^7^ and vocabulary^2^ tend to be preserved. Importantly, cognitive functions rarely operate in isolation; rather, they interact dynamically. For instance, higher working memory capacity and vocabulary can support episodic memory retrieval in older adults^8–10^. Beyond age-related changes such as synaptic alterations^11,12^, most cognitively unimpaired older adults harbor^13,14^ tau tangles within the medial temporal lobe^15,16^, one hallmark of Alzheimer’s disease. Some cognitively normal older adults also harbor neocortical amyloid-beta (Aβ) plaques^17,18^, which is often associated with unfavourable cognitive trajectories^19^, rendering the detection of early subtle cognitive decline an important aim.

One of the major risk factors for sporadic Alzheimer’s disease is the Apolipoprotein-E4 (*APOE4*) genotype^20,21^, which is strongly correlated with Aβ accumulation^22,23^. While some studies report subtle early cognitive deficits in *APOE4* carriers already in midlife^24,25^, most studies did not find *APOE*-related cognitive or neural differences in middle-aged adults (for reviews, see ^26,27^). In cognitively unimpaired older *APOE4* carriers, deficits in visual mnemonic discrimination^28^ and in episodic memory retrieval of words^29^ have been reported, whereas another study reported no effects of *APOE4* on object-location binding^30^. In older adults with subjective cognitive decline (SCD), older *APOE4* carriers showed a steeper decline in general cognition compared to cognitively unimpaired *APOE4* carriers^31^. These mixed results underscore the importance of identifying subtle early cognitive decline in vulnerable individuals before a decline in general cognition becomes apparent and further accelerates. Furthermore, in the era of Alzheimer’s disease treatment^32–34^, early detection of individuals in need of monitoring and medical intervention is key. An important question to answer is therefore how to detect early subtle cognitive deficits to identify these individuals who are still cognitively unimpaired in standard neuropsychological testing.

Theories regarding intact cognition and brain function despite the loss of neural resources, e.g., due to Alzheimer’s pathology, are gathered under the umbrella term of resilience^35,36^. One mechanism of resilience is functional compensation: the short-term recruitment of additional neural resources in response to higher task demands. Task demand influences cognitive performance, with more demanding task conditions creating a greater cognitive challenge that needs to be met with the available neural resources^37–40^. During a cognitive task, short-term changes in brain network activity, e.g., more recruitment of domain-general cognitive control brain networks^41,42^, may be required to meet the current task demands. When fewer neural resources are available due to age– or Alzheimer’s disease-related processes, more effort is needed to meet task demands at a lower demand level^37,43^ and a failure of compensatory mechanisms could occur earlier^44^. For an overview of theories and a discussion of compensatory vs. detrimental effects, see ^45^. A resulting question is, how to empirically investigate these theoretically proposed mechanisms of compensation related to episodic memory retrieval task demand.

One experimental approach is to gradually increase task demands across multiple levels to create a stepwise increase in cognitive challenge^46–49^. Few studies to date have, however, used a manipulation of retrieval-task demand to investigate episodic memory^50^. Furthermore, aging studies often do not consider Alzheimer’s disease risk factors and pathology in cognitively unimpaired older adults, which is crucial for understanding processes related to aging^3,51,52^. To address this research gap, we adapted a behavioural task previously used for immediate retrieval^48,53,54^ to implement a manipulation of task demand during delayed retrieval. Within the EMCOMP (Episodic Memory & COMPensation) study, we first assessed retrieval performance across five recognition and two recall demand levels in young adults and cognitively unimpaired older adults. Second, we assessed the influence of the *APOE4* genotype and plasma-based Alzheimer’s pathology burden on retrieval performance in older adults. Third, we assessed the relation to performance in established cognitive tests in older adults.

We hypothesized that i) retrieval accuracy declines with increasing task demand, ii) older compared to younger adults, *APOE4* carriers compared to non-carriers, and individuals with more Alzheimer’s pathology show lower recognition accuracy, recall less correct information, and show a steeper decline in performance with higher task demand, and iii) older adults’ confidence in their response follows a U-shape, i.e. an initial decrease in confidence (in the correct response) as demand increases, followed by an increase in confidence (in the incorrect response) as demand increases further. In individuals with the *APOE4* genotype and higher Alzheimer’s pathology burden, these effects become apparent even at lower demand levels.

## 2. Methods

### 2.1. Sample

We collected data from 37 young and 63 older adults. All participants were native German speakers, had normal or corrected to normal vision, and reported no history of neurological or mental disorders including dyslexia. All participants were required to reach at least 80% accuracy in the encoding cover task (see section 2.2.2 “Encoding session”) to ensure attention on the task.

Young adults were primarily recruited via online advertisement and were predominantly students at the Otto-von-Guericke University Magdeburg and the University of Applied Sciences Magdeburg-Stendal. Older adults were recruited from the participant pool of the Z03 project which is part of the Collaborative Research Centre (CRC) 1436 “Neural Resources of Cognition” (see ^55^ for details).

In short, older participants were over the age of 60 and cognitively unimpaired with scores within their age range of the CERAD-Plus (Consortium to Establish a Registry for Alzheimer’s Disease) Neuropsychological Assessment Battery. Additionally, they had no major age-related illnesses. After excluding 8 participants (2 young adults did not meet the inclusion criteria, 4 older adults misunderstood the task, 2 older adults did not reach the threshold for the encoding cover task), the final sample consisted of 35 young (24±3years, 17 female) and 57 older adults (72±0years, 31 female, 14 *APOE4* carriers). *APOE4* carriers and non-carriers did not differ significantly in demographic variables. Details and additional information can be found in Table 1.

**Table 1.** Description of the sample.

| Age group | N | Age in years | Female sex | Education in years | Vocabulary score | Working memory capacity | *APOE4* carrier | Alzheimer's disease pathology (AT-Term) |
| --- | --- | --- | --- | --- | --- | --- | --- | --- |
| Young adults | 35 | 23.8 (2.9)<br>18 - 30 | 17 (49%) | 16.5 (2.2) | 99.0 (4.7) | 0.9 (0.5) | - | - |
| Older adults | 57 | 72.0 (7.2)<br>62 - 89 | 31 (54%) | 15.6 (2.4) | 112.0 (9.3) | 0.6 (0.8) | 14 (25%) | 1.45 (1.01) |

Mean (standard deviation) or number (percentage) of participants. Age denotes age at baseline, age range is additionally provided. All participants identified as either female or male. Three *APOE4* carriers were homozygous. For the vocabulary score, Mehrfachwahl-Wortschatz-Intelligenztest (MWT-B) normative values were used. Working memory capacity was assessed using a change detection task. For Alzheimer's disease pathology, the AT-Term $1 / (A\beta_{1-42} / A\beta_{1-40}) * p\text{-tau}_{217}$ was used.

In our preregistration^56^, we planned to recruit 35 young and 35 older adults for age-group comparisons for a Bachelor Thesis, resulting in a total of 70 participants. We estimated this required sample size using G*Power for a multiple linear regression with recall performance as the dependent variable and age group, recall demand, sex, vocabulary score, and working memory score as predictors. Assuming a medium-to-large effect size (f² = 0.20), an α-level of 0.05, and a desired power of 0.80, the analysis yielded a required sample size of 70 participants. In addition, we predetermined to continue collecting and processing data from older adults for the analysis of *APOE4* group and Alzheimer’s pathology until October 2025. This pre-specified end point was based on internal personnel resources.

All study procedures and experimental protocols were approved by the Ethics Committee of the Medical Faculty, Otto-von-Guericke University Magdeburg (200/19) and were carried out in accordance with the Declaration of Helsinki. All participants provided written informed consent.

### 2.2. Episodic memory retrieval task-demand manipulation

#### 2.2.1. Stimuli

The experiment included three main sessions: encoding, recognition, and recall. For each participant an individual encoding item (i.e., sentence) list was created with the same 75 sentences in a randomized order. Further, one encoding-practice list for all participants was created using five sentences. For the recognition task, a list of 93 sentences was constructed including sentences of all five recognition-demand levels from easy to hard: 18 completely novel foils, 21 “old” target sentences (were presented during encoding), 18 easy lures, 18 medium lures and 18 hard lures. The allocation of original sentences to recognition demand level was semi-randomized over all three sentence types. Recognition demand levels were based on previous work^57,58^. Further, a recognition-practice list was created with retrieval-demand manipulation of the encoding practice sentences. Stimuli are openly available on the OSF (https://doi.org/10.17605/OSF.IO/E4MC9). In the following, the construction of base and lure items is described.

Of all 93 written German sentences, 75 sentences were first seen at encoding, the remaining 18 were first seen at recognition as novel foils. All sentences were similar in length (approximately 13 words) and included unique content and words. The construction of the stimuli was based on previous work, where meaningful 10-word sentences were used to investigate syntactic working memory load and immediate recall^46,48^. In this study, the sentences from DeCaro and colleagues^48^ were translated from English into German. Then, the sentences were extended with additional information, including social and spatial relations, to provide both gist-level and detailed content for later retrieval. All sentences underwent an unpublished stimuli rating in which 8 young and 6 older adults (N = 14) read the sentences without time restrictions and assessed them on a 9-point scale (from –4 to 4) in the following categories: emotion, schema congruency, comprehensibility, word frequency and memorability. This rating was conducted to ensure that the sentences were approximately similar in the respective categories when no challenge due to a time restriction was present. The rating resulted in five sentences being conspicuous in terms of schema congruency, so they were altered to be more congruent.

Three different sentence types with a total of 31 subject-relative sentences with a long agent-action gap (subject-long), 31 object-relative sentences with a short gap (object-short) and 31 object-relative sentences with a long gap (object-long) were used (for details, see ^48^; for examples, see Table 2). Object-relative syntactic structures are cognitively more challenging compared to syntactically simpler subject-relative structures^48^. This design was implemented for future use of the task in the MRI scanner, where different levels of encoding-demand will be investigated in relation to brain activation during encoding. In this behavioural pre-study, the encoding session was only encompassing a cover task (see section 2.2.2 “Encoding session”). Use of names, sex, and plural form was balanced across the three sentence types. Sentence types were balanced across encoding, “old” target, and novel items.

**Table 2.** Examples of base items.

| Sentence type | Example |
| --- | --- |
| Subject-long | Die <u>Mütter</u> aus dem dritten Stock, die Kai zum Fußballspiel <u>einladen</u>, sind sympathisch.<br>The <u>mothers</u> from the third floor who <u>invite</u> Kai to the football match are likeable. |
| Object-short | Der junge Elektriker aus der Nachbargemeinde, dem <u>Olivia</u> <u>hilft</u>, ist endlich selbständig.<br>The young electrician from the neighboring community, whom <u>Olivia</u> <u>helps</u>, is finally self-employed. |
| Object-long | Die Enkeltochter, der <u>Opa Herbert</u> mit Schmerzen im unteren Rücken <u>vorliest</u>, ist schläfrig.<br>The granddaughter, whom <u>Grandpa Herbert</u> with pain in the lower back <u>reads</u> to, is sleepy. |

One example for each of the three sentence types of base stimuli in German with a literal translation in English below. “Object-long” means an object-relative sentence with a long agent-action gap. For agent-action gap visualization, the agent and the action of the sentence are underlined. Cognitive challenge is increasing from subject-long to object-short to object-long syntaxes <sup>48</sup>.

For the recognition task, 54 lures (three recognition-demand levels; 18 stimuli per level) were created. As lure items, encoded stimuli were altered resulting in novel items with varying levels of similarity to the originally encoded items. For the easy lure level, two words were replaced by non-synonymous words (e.g., “apron” replaced by “wig” and “diligently” replaced by “lazily”; see Table 3) which changed the meaning of the original sentence. For medium lure level, only one word was replaced by a non-synonymous word (e.g., “colleague” replaced by “trainee”), that is however part of the same word category so that the content only changed slightly. For the hard lure level, one word was exchanged with a synonym, so the meaning of the sentence remained the same (e.g., “black” replaced by “dark”). The full example sentences are provided in Table 3. To check synonyms, the online Oxford Languages English Dictionary^59^ was used. To keep the word frequency of the exchanged words consistent, the online German Dictionary “DWDS – Digitales Wörterbuch der Deutschen Sprache”^60^ was used.

**Table 3.**
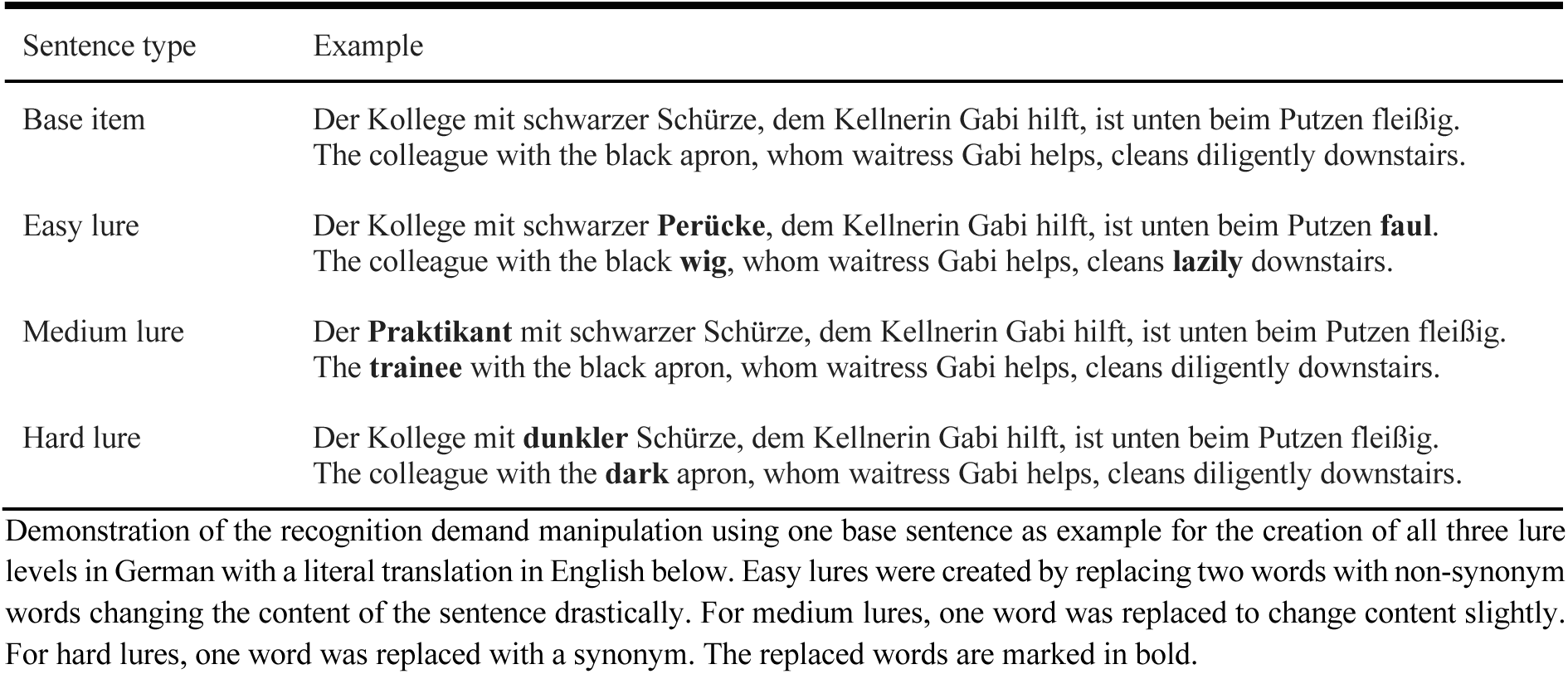
Examples of lure items.

#### 2.2.2. Encoding session

The encoding session was conducted using Psychopy version 2023.2.3^61^. The instruction, the items and the fixation cross were presented in white (RGB: 255,255,255) on a gray background (RGB: 128,128,128). In the intentional encoding session, each participant viewed all 75 target items. Participants were instructed to read each sentence, try to memorize it and indicate via keypress whether the first person mentioned was female or male. The female/male decision task was included as cover task to ensure that the respective sentence was processed by the participant. However, it was emphasized in the instruction that the focus of the encoding task was to memorize the sentences. Each trial of the encoding task (see Figure 2) started with a fixation cross that was randomly displayed for two to six seconds. The randomized presentation time of the fixation cross was set to minimize habituation effects and used in light of the planned implementations of the task in the fMRI scanner. After the fixation cross, a sentence was presented for seven seconds in which the participants had time to read the sentence thoroughly and remember as much as possible. Seven seconds for encoding were determined in our preregistration based on the self-paced reading speed of young adults (400 ms/word) and older adults (700 ms/word)^62^. For a 13-word sentence, this corresponds to approximately five and nine seconds, respectively. As the aim of the study was to create a cognitive challenge, especially in older adults, the mean combined self-paced reading speed of both age groups was used, which resulted in seven seconds. The item presentation was followed by another randomized presentation of the fixation cross, for two to six seconds, followed by the decision task to indicate the gender of the first person mentioned. Participants responded via keypress (F = female, J = male), within a response window of two seconds. One trial took about 15 seconds. The encoding task including the practice part of five trials took about 25 minutes.

**Figure 2.**
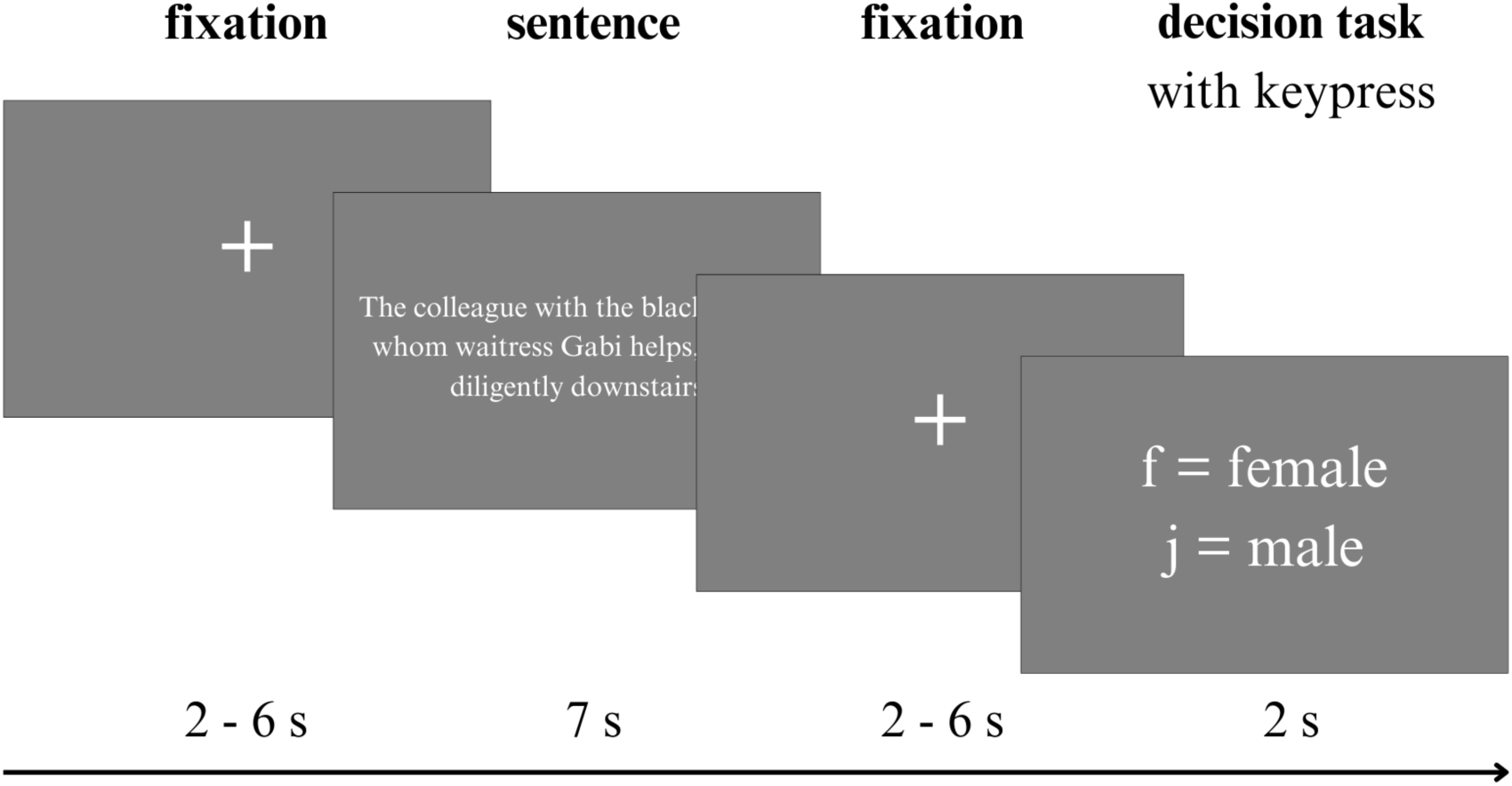
Procedure of one trial in the encoding task. This figure shows the procedure of one trial in the encoding task. First, a fixation cross was presented, then the target item (sentence) was presented, followed again by a fixation cross of variable duration. Then the decision task followed to indicate the gender of the first person mentioned. Answers were given via key press.

#### 2.2.3. Recognition session

The recognition session started 15 minutes after the encoding session ended. Within these 15 minutes, participants watched a silent nature documentary. The same Psychopy setup was used as for the encoding session. The main task was the identification of “old” target sentences that participants had encoded before, in contrast to lures and novel foils. Participants viewed 93 stimuli, including 21 “old” target sentences, 18 novel foils and 54 lure sentences (18 in each lure level). Participants were instructed to read the displayed sentence and decide whether it was “old” (already seen during encoding) or “new” (lure or novel foil). Additionally, they should indicate their confidence in their categorization of the sentence. Both categorization and confidence were reported in a confidence rating via keypress. The structure of the trials in the recognition task (see Figure 3) was similar to that of the encoding task. Each trial started with the presentation of the fixation cross for a randomized period of two to six seconds. Then, a sentence was displayed for seven seconds. After another presentation of the fixation cross, the confidence rating followed. Participants again had a time window of two seconds to give their answer via keypress. They were instructed to press 1 if they were sure they had read the sentence in the encoding task before (= sure old), press 2 if they were unsure but thought the sentence was probably old (= probably old). The same principle applied if they thought the sentence was new: 8 = probably new; 9 = sure new. One trial lasted for about 15 seconds, followed by the next trial. Similar to the encoding task, there was a practice part before the main recognition task started. The recognition task took about 25 minutes.

**Figure 3.**
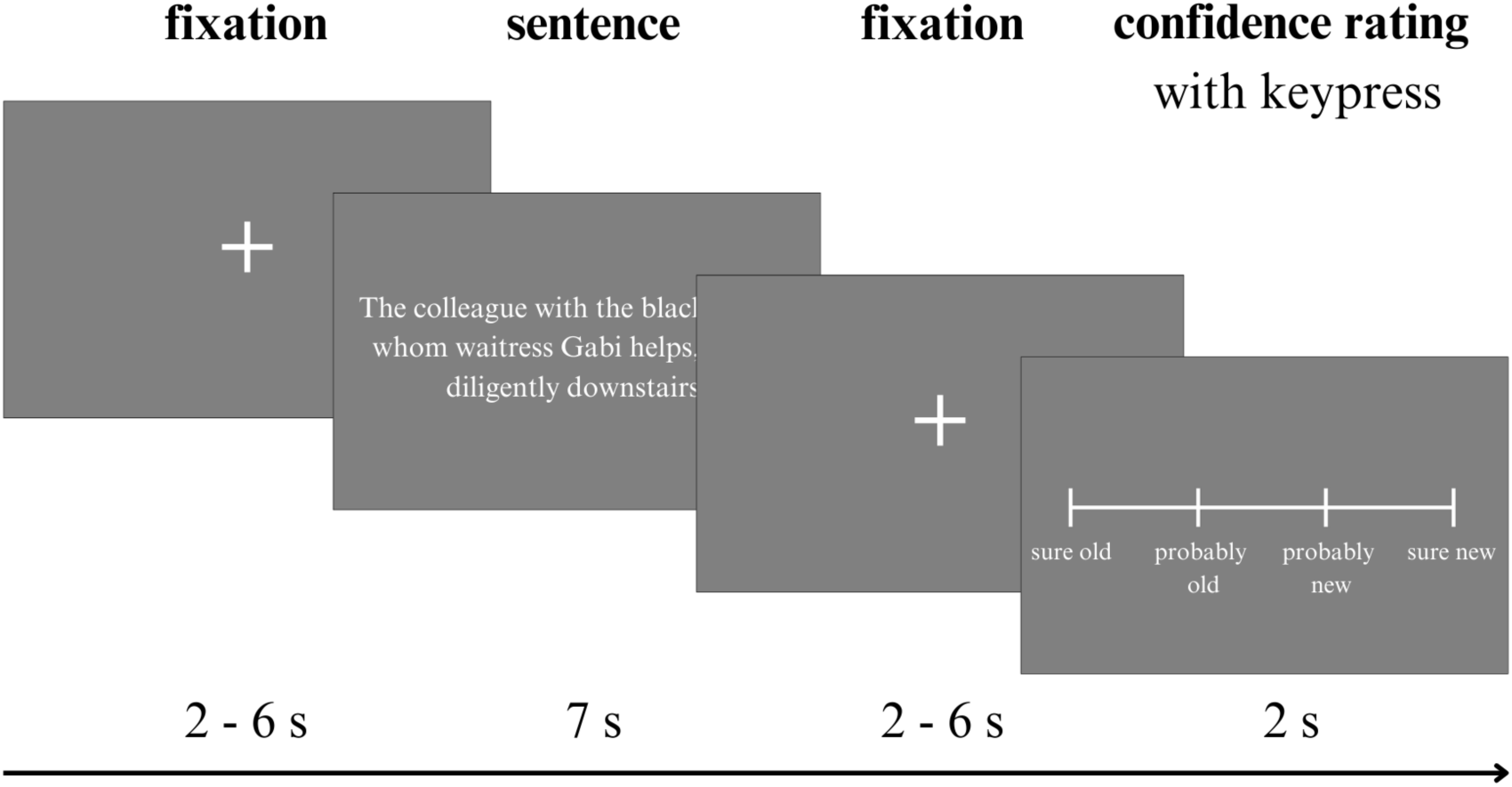
Procedure of one trial in the recognition task. This figure shows the procedure of one trial in the recognition task. First, a fixation cross was presented, then the demand manipulated target item was presented (new or old or lure level), before the confidence rating followed, after another fixation cross. Answers were given via key press.

#### 2.2.4. Recall session

The recall session was divided into two parts: free recall (approximately 90 minutes after start of encoding) and cued recall (approximately 100 minutes after start of encoding). In the free recall task, participants were prompted with the question, “Which sentences do you remember from the first (encoding) experiment?”. They were encouraged to reproduce everything they remembered, full sentences and phrases or fragments of the sentences were possible. When they had recalled everything they knew, the cued recall followed. The instructor read the first half of every sentence (6 words) out loud as a cue. If participants remembered anything about the second half, they were supposed to complete the sentence. Both recall types were audio-recorded to simplify the evaluation and to ensure that no information was lost. Afterwards the audio was transferred into text and scored with a recall scoring system (see Supplementary Methods S1 and Table S1). Then, a relative recall score was computed, by dividing each participant’s obtained recall score by the maximum possible score, assuming complete recall.

### 2.3. Cognitive testing

For measuring the participants’ vocabulary, the B Version of the German Mehrfachwahl-Wortschatz-Intelligenztest (MWT-B)^63^ (translated: “multiple choice vocabulary intelligence test”) was used. The MWT-B is used to measure the level of crystalline intelligence. Participants are instructed to find the existing German word among four other invented non-existing similar words. The words existing in the German language originate from colloquial, educational or scientific contexts and become more specific and rare with each row. In total there are 37 rows with five words each. Z-values were derived using the test manual.

A computer-based change detection task was carried out to measure visual working memory capacity. In this experiment, an open-source version^64^ was used. In the change detection paradigm, participants first see a fixation cross for 1 second, an array of six visual items (squares) of different colors for 0.5 second, another fixation cross as delay phase for 1 second, and then a single test item (colored square) as probe until keypress. Participants had to decide via keypress whether the position and color of the probe is similar to the position and color of one of the presented items^65^. In this study, the visual working memory capacity K was calculated using a standard formula^66,67^: K=N× (H − FA), where K is the capacity estimate, N is the set-size, H is hit rate (proportion of correct responses in change trials), and FA is the false alarm rate (proportion of incorrect responses in no-change trials).

Scores from neuropsychological assessment of older adults included tests focussing on episodic memory and other cognitive functions. For details, see ^55^. Regarding episodic memory, the Verbal Learning and Memory Test (VLMT)^68^ with 15 words, and the Rey Complex Figure Test and Recognition Trial (RCFT)^69^ with one complex figure were used for delayed free recall and recognition. Further, Logical Memory (LM) II of the Wechsler Memory Scale (WMS)^70^ was used for delayed free recall of stories; a German picture version of the Free and Cued Selective Reminding Test (FCSRT)^71^ provided a sum score of free and cued immediate recall.

Regarding further cognitive functions, the Symbol Digit Modalities Test (SDMT)^72^ was used to assess processing speed. The German Regensburger Wortflüssigkeitstest (RWT)^73^, which includes semantic and phonemic verbal fluency tasks, was used to assess executive functioning. The CERAD-Plus Neuropsychological Assessment Battery^74^ was used to assess general cognition with a composite score including: Mini-Mental State Examination (MMSE), verbal and phonemic fluency, a short form of the Boston Naming Test, 10-word list learning and delayed recall and recognition, constructional praxis of figures and delayed recall, and the Trail Making Test (TMT) version A and B.

### 2.4. Assessment of Alzheimer’s disease plasma biomarkers

Plasma Aβ_1–40_, Aβ_1–42_, and p-tau_217_ were determined via Lumipulse assays for older adults. For details, see ^55^. To investigate a composite measure of Alzheimer’s disease pathology, the AT-Term = 1/ (Aβ_1-42_/Aβ_1-40_) * p-tau_217_ was used.

### 2.5. *APOE* genotyping

All included older participants were genotyped for *APOE* via a polymerase chain reaction-based approach followed by restriction fragment length polymorphism analysis. For details, see ^55^. *APOE4* carriers were defined as individuals with one or two *APOE4* alleles. *APOE4* non-carriers were defined as individuals without an *APOE4* allele.

### 2.6. Study design

A preregistration of *a priori* hypotheses and methods was uploaded on the Open Science Framework (OSF)^56^ before data collection was completed and before data was compiled and accessed. For investigating how recognition demand (five levels: novel items, “old” target items, easy lure, medium lure, hard lure) and age group (two levels: young adults, older adults), or *APOE4* group (two levels: carrier, non-carrier) and the AT-Term (continuous measure) in older adults only, influence recognition performance, the dependent variables were accuracy and confidence. In analysis including only older adults, age was treated as a continuous predictor. Recognition accuracy refers to the number of correct trials relative to all trials per demand level, and confidence refers to the subjective assessment of one’s own success in categorizing a sentence correctly. For investigating effects on recall performance, recall demand (two levels: gist, detail) was examined in relation to age group or *APOE4* group and Alzheimer’s pathology. The dependent variables were recall score and the recall error rate (swaps and add ons). They all resulted from the recall scoring system (see Supplementary Methods S1 and Table S1).

### 2.7. Statistical analysis

Statistical analysis was conducted using R Version 4.3.1 (RStudio Team, Boston, MA, USA) implemented in R studio (Version 2024.12.1). The R code used for analyses is publicly available (https://github.com/fislarissa/EMCOMP_demand). Multiple linear regression and linear mixed-effects models (LMM) with a random intercept for participant were used. In all models, sex, vocabulary score and working memory capacity were included as covariates as predefined in the preregistration based on a previous study^10^. Education was not used as a covariate, as a further planned analysis aims at investigating education as a proxy for cognitive reserve. The Z03 measurements in older adults took place before the EMCOMP measurements (median 15 months, range 3 to 38 months). Including the individual time difference in the models did not change the results. The statistical level of significance was set at *p* < 0.05 for all analyses. Regarding model requirements, it was ensured that the assumption of homoscedasticity was met using the Breusch-Pagan test and that no multicollinearity was present with a variance inflation factor (VIF) below 5. Further, the assumption of a normal distribution of residuals was assessed using quantile-quantile (QQ) plots. Multiple comparisons were controlled using False-discovery-rate (FDR) correction according to the Benjamini-Hochberg procedure.

First, we investigated the influence of recognition-demand level and the interaction of recognition-demand level by age group on recognition accuracy in an LMM (*Accuracy ∼ Recognition-demand level (linear) * Age group + Sex + Vocabulary + Working memory capacity)*. We then investigated the influence thereof on recognition confidence, including a U-shape association of confidence across demand levels (*Confidence ∼ Recognition-demand level (linear) * Age group + Recognition-demand level (U-shape) * Age group + Sex + Vocabulary + Working memory capacity)*, and further investigated the influence of accuracy on recognition confidence (*Confidence ∼ Recognition-demand level (linear) * Age group * Accuracy + Sex + Vocabulary + Working memory capacity)*.

Then we investigated the influence of recall-demand level and the interaction of recall-demand level by age group on recall score (*Recall score ∼ Recall-demand level (linear) * Age group + Sex + Vocabulary + Working memory capacity)* and recall error rate (*Recall error rate ∼ Recall-demand level (linear) * Age group + Sex + Vocabulary + Working memory capacity).* As the residuals were not normally distributed, a log-transformation was applied to the error rates. For interpretation of the results on the original scale, the regression coefficients (β) were additionally exponentiated (exp(β)).

Second, only in older adults, we repeated the described models with *APOE4* group instead of age group. Additional predictors were age at baseline and the AT-Term. Exploratively, we repeated these models to investigate VLMT and RCFT recognition performance instead of EMCOMP performance.

Third, only in older adults, we conducted correlation analysis for i) recognition accuracy, using the EMCOMP corrected hit rate (hits (categorizing “old” target items correctly as old) – false alarms (categorizing novel items incorrectly as old)) and corrected hit rate for the episodic memory tests VLMT and RCFT; ii) recall, using the EMCOMP recall score across both demand levels and the delayed recall scores of the episodic memory tests VLMT, RCFT, WMS, and immediate recall score of the FCSRT; iii) established cognitive tests, using the EMCOMP corrected hit rate and EMCOMP recall score across both demand levels and test scores for processing speed (SDMT), executive functioning (RWT) and general cognition (CERAD).

## 3. Results

### 3.1. Retrieval performance in young and older adults

First, we investigated the effect of recognition-demand level and the interaction of recognition-demand level by age group on recognition accuracy (see Table 4 for descriptive values, Figure 4a, and Supplementary Table S2). We included sex, vocabulary, and working memory capacity as covariates, as in all following LMMs. The model revealed that in both age groups, accuracy decreased with increasing recognition-task demand (β=-0.63 [95% CI –0.73, –0.53], t=-12.71, *p*<0.001) and accuracy was highest for identifying novel foils and lowest for discriminating hard lures. Older adults performed worse than young adults across all recognition-demand levels (β=-0.35 [95% CI –0.52, –0.28], t=-3.99, *p*<0.001). In addition, there was an interaction effect of recognition demand by age group (β=-0.13 [95% CI –0.25, –0.00], t=-2.01, *p*=0.045, Cohen’s f²=0.01), with older adults’ accuracy showing a steeper decline with increasing recognition-task demand.

**Figure 4.**
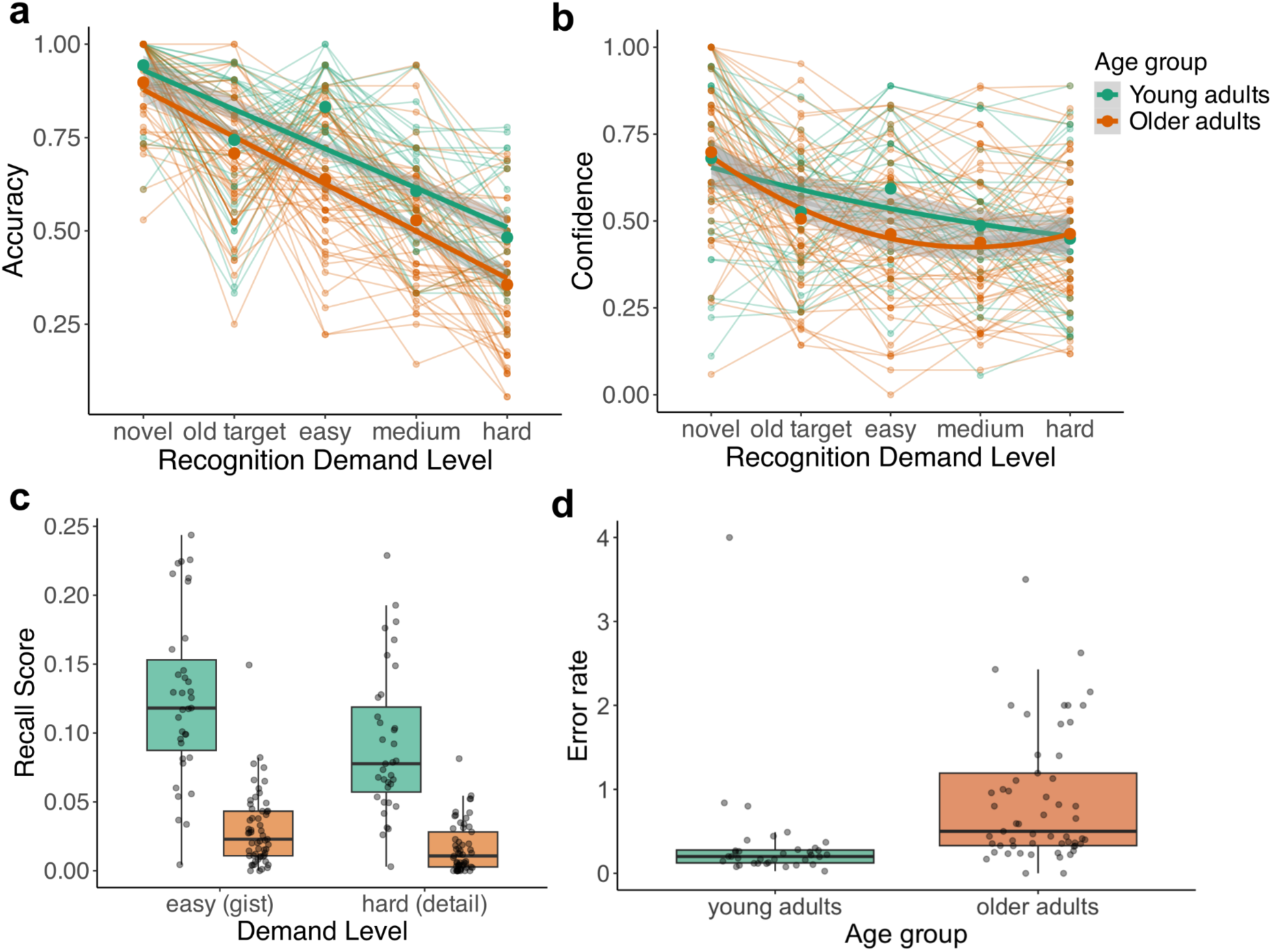
Recognition-task performance for young and older adults. a) Accuracy across recognition-task demand levels for young (green lines) and older adults (orange lines). Presented are group means (bold dots), smoothed group mean trajectories (bold lines) and individual trajectories (fine lines). b) Confidence across recognition-task demand levels for young (green lines) and older adults (orange lines). Presented are group means (bold dots), smoothed group mean trajectories (bold lines) and individual trajectories (fine lines). c) Recall score for both recall-demand level (gist and detail) ranging from 0 to 1. A score of 0.10 means that 10% of the information the participant was exposed to was recalled. The median is shown as bold black horizontal line and the 25th and 75th percentile as black horizontal lines below and above. d) Recall error rate averaged across recall-demand level (gist and detail). The median is shown as bold black horizontal line and the 25th and 75th percentile as black horizontal lines below and above.

**Table 4.** Accuracy and confidence of young and older adults.

|  | Accuracy |  | Confidence |  |
| --- | --- | --- | --- | --- |
| Recognition demand | Young adults<br>(N = 35) | Older adults<br>(N = 57) | Young adults<br>(N = 35) | Older adults<br>(N = 57) |
| Novel foils | 0.94 (0.10) | 0.90 (0.11) | 0.68 (0.25) | 0.70 (0.22) |
| “Old” target items | 0.74 (0.18) | 0.71 (0.17) | 0.52 (0.16) | 0.51 (0.20) |
| Easy lure items | 0.83 (0.10) | 0.64 (0.18) | 0.59 (0.18) | 0.46 (0.20) |
| Medium lure items | 0.61 (0.15) | 0.53 (0.17) | 0.49 (0.17) | 0.44 (0.21) |
| Hard lure items | 0.48 (0.15) | 0.36 (0.17) | 0.45 (0.21) | 0.46 (0.19) |

Mean (standard deviation) for accuracy and confidence of 35 young and 57 older adults. Accuracy is the number of correct trials relative to all trials of the respective recognition demand level. Confidence is the number of trials where participants indicated to be confident in their response relative to all trials of the respective recognition demand level.

We then investigated the effect of recognition-demand level and the interaction of recognition-demand level by age group on recognition confidence (see Table 4 for descriptive values, Figure 4b, and Supplementary Table S3). Here we additionally assessed a U-shaped relationship additionally to the linear relationship across recognition-demand levels. The model revealed that confidence decreased with increasing recognition-task demand in a linear (β=-0.32 [95% CI –0.42, –0.22], t=-6.30, *p*<0.001) manner. Older adults were less confident than young adults in all recognition-task demand levels (β=-0.43 [95% CI –0.84, –0.01], t=-2.03, *p*=0.044), however, there was an interaction effect of recognition-task demand (U-shape) by age group (β=0.24 [95% CI 0.09, 0.39], t=3.11, *p*=0.002, Cohen’s f²=0.01), with older adults’ confidence showing a U-shape pattern of confidence across demand levels.

Additionally, we investigated the influence of accuracy and the interaction of recognition-demand level by age group by accuracy on recognition confidence (see Table 4 for descriptive values, Supplementary Figure S1 and S2, and Supplementary Table S4). The model revealed that confidence was higher for correct compared to incorrect responses (β=0.59 [95% CI 0.51, 0.67], t=14.35, *p*<0.001). There was an interaction effect of recognition demand by accuracy (β=-0.21 [95% CI –0.29, –0.13], t=-5.01, *p*<0.001, Cohen’s f²=0.10), with confidence for correct responses decreasing while confidence for incorrect responses increasing with higher demand level. Further, there was an interaction effect of age group by accuracy (β=-0.32 [95% CI –0.42, 0.22], t=-6.19, *p*<0.001, Cohen’s f²=0.04), with older adults showing lower confidence compared to young adults for correct responses. The three-way interaction of recognition-demand level by age group by accuracy with older adults showing a descriptively steeper decline in confidence across recognition-task demand compared to young adults for correct responses was not significant (β=-0.10 [95% CI –0.20, –0.00], t=-1.91, *p*=0.056).

Further, we investigated the effect of recall-demand level and the interaction of recall-demand level by age group on recall performance (see Figure 4c, and Supplementary Table S5) based on a recall scoring system (see Supplementary Methods S1 and Table S1). A mean recall score of cued and free recall was used (correlation of cued and free recall: r = 0.76 [95% CI 0.64, 0.85], t=9.53, *p*<0.001). The model revealed that performance decreased with increasing recall-task demand (β=-0.29 [95% CI –0.34, –0.24], t=-12.25, *p*<0.001). Older adults recalled less than young adults for both recall-demand levels (β=-1.46 [95% CI –1.82, –1.09], t=-7.86, *p*<0.001), however, there was an interaction effect of recall-demand level by age group (β=0.17 [95% CI 0.11, 0.23], t=5.78, *p*<0.001, Cohen’s f²=0.02), with young adults’ performance showing a steeper decline across demand levels. Caution should be taken regarding this interaction, as the recall score for older adults for gist (mean=0.030, SD=0.027) and detail (mean=0.016, SD=0.018) is very low compared to young adults’ scores for gist (mean=0.126, SD=0.06) and detail (mean=0.092, SD=0.053). A score of 0.092 means that 9.2% of the information the participant was exposed to was recalled.

Additionally, we investigated the influence of age group on recall error (see Figure 4d, and Supplementary Table S6). The model revealed that the recall error was higher for older compared to young adults (β=0.34 [95% CI 0.13, 0.55], t=3.22, *p*=0.002), with older adults’ showing on average 2.5 times the error rate of young adults.

Taken together, young adults showed higher recognition accuracy and confidence as well as a higher recall score and lower recall error. Older adults’ accuracy declined more steeply than young adults’s accuracy with increasing recognition-task demand.

### 3.2. Retrieval performance in older adults related to *APOE4* genotype and Alzheimer’s pathology

Second, only in older adults, we investigated the influence of recognition-demand level, the interaction of recognition-demand level by *APOE4* group, and the effect of the plasma-based Alzheimer’s pathology on recognition accuracy (see Table 5 for descriptive values, Figure 5a, and Supplementary Table S7). We included age at baseline, sex, vocabulary, and working memory capacity in all following LMMs. The model revealed that accuracy decreased with increasing recognition-task demand (β=-0.70 [95% CI –0.78, –0.61], t=-15.65, *p*<0.001). There was no main effect of *APOE4* group (β=-0.09 [95% CI –0.28, 0.10], t=-0.92, *p*=0.360) or the AT-Term (β=0.05 [95% CI –0.03, 0.14], t=1.16, *p*=0.245). However, there was an interaction effect of recognition-demand level by *APOE4* group (β=-0.18 [95% CI –0.36, – 0.01], t=-2.06, *p*=0.041, Cohen’s f²=0.02), with *APOE4* carriers showing a steeper decline in accuracy with higher recognition-task demand.

**Figure 5.**
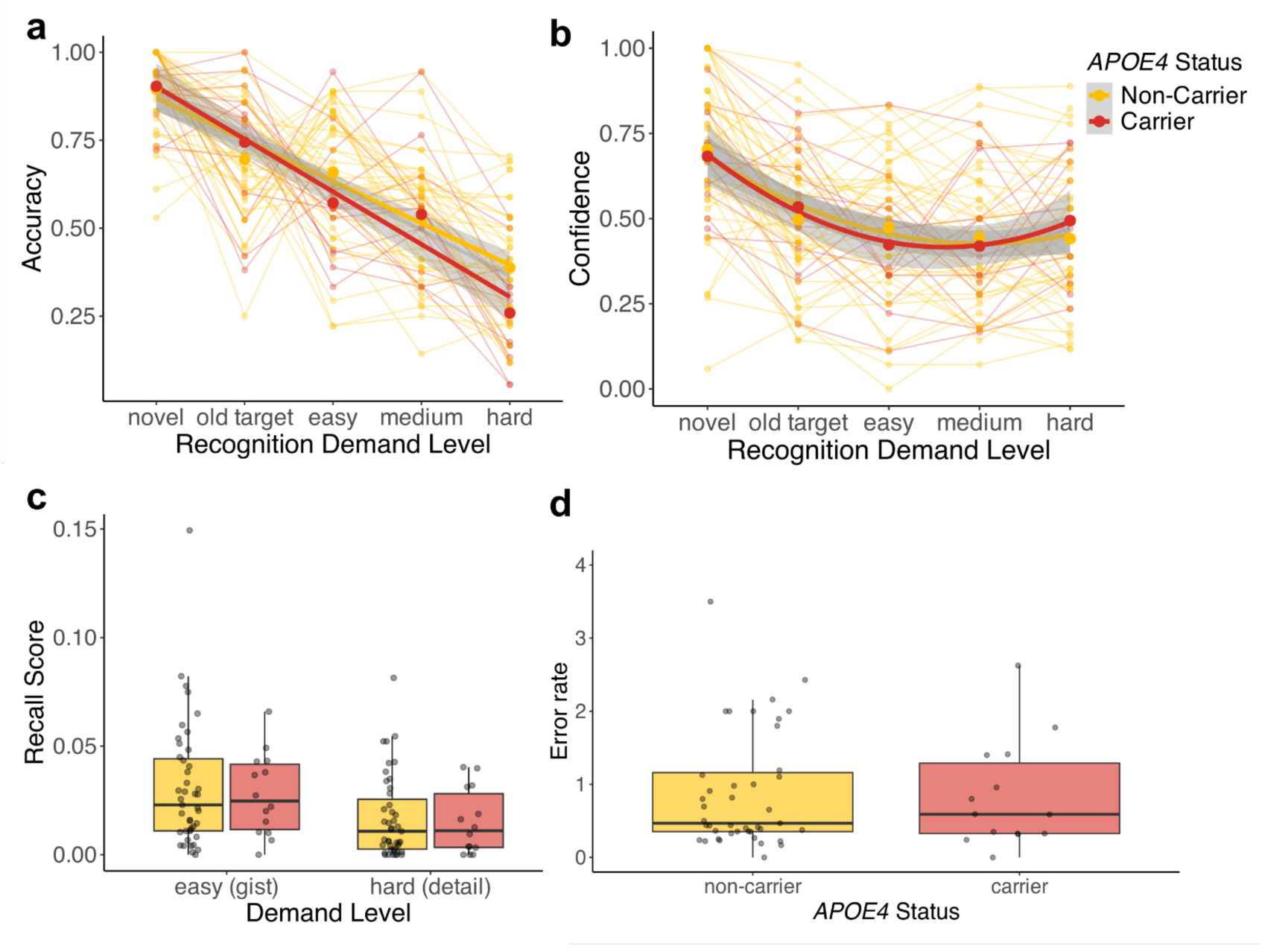
Recognition-task performance for older *APOE4* carriers and non-carriers. a) Accuracy across recognition-task demand levels for young (green lines) and older adults (orange lines). Presented are group means (bold dots), smoothed group mean trajectories (bold lines) and individual trajectories (fine lines). b) Confidence across recognition-task demand levels for young (green lines) and older adults (orange lines). Presented are group means (bold dots), smoothed group mean trajectories (bold lines) and individual trajectories (fine lines). c) Recall score for both recall-demand level (gist and detail). The median is shown as bold black horizontal line and the 25th and 75th percentile as black horizontal lines below and above. d) Recall error rate for both recall-demand level (gist and detail). The median is shown as bold black horizontal line and the 25th and 75th percentile as black horizontal lines below and above.

**Table 5.**
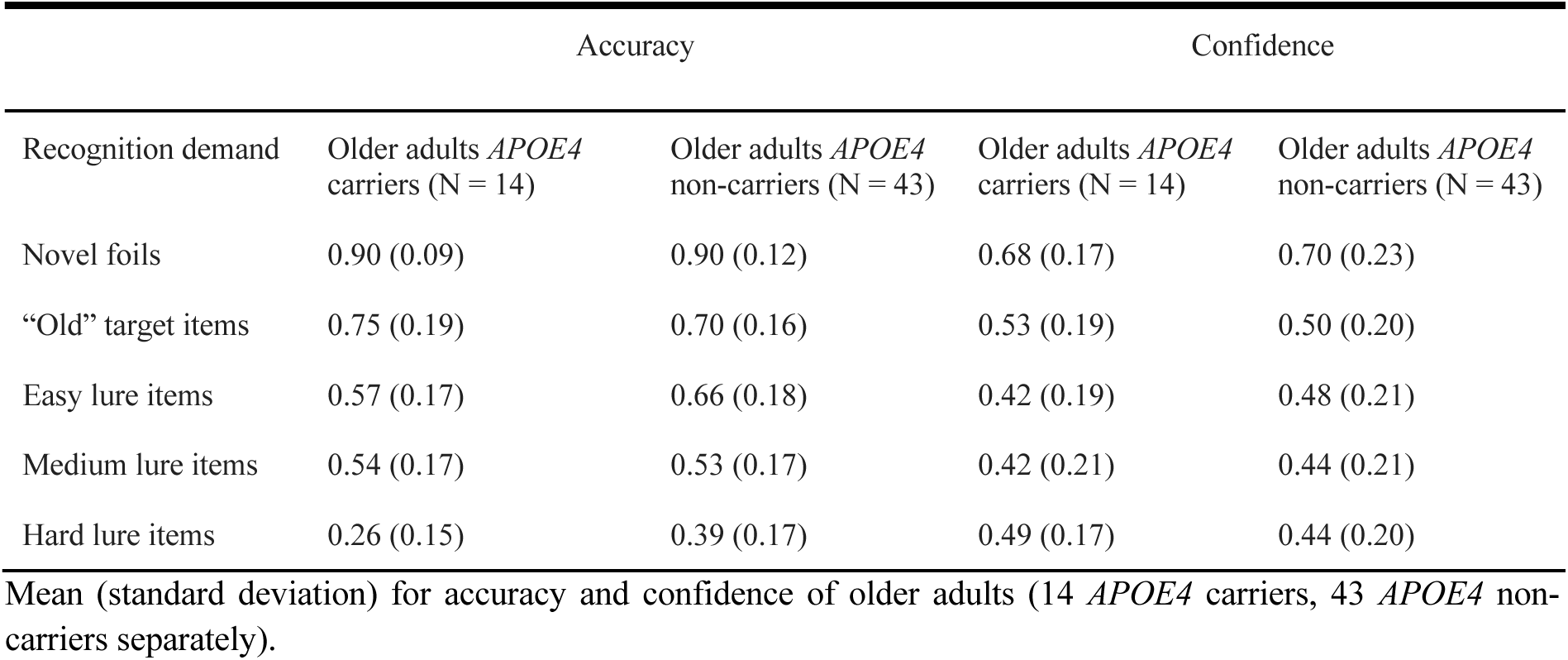
Accuracy and confidence of older *APOE4* carriers and non-carriers.

| Recognition demand | Accuracy |  | Confidence |  |
| --- | --- | --- | --- | --- |
|  | Older adults *APOE4* carriers (N = 14) | Older adults *APOE4* non-carriers (N = 43) | Older adults *APOE4* carriers (N = 14) | Older adults *APOE4* non-carriers (N = 43) |
| Novel foils | 0.90 (0.09) | 0.90 (0.12) | 0.68 (0.17) | 0.70 (0.23) |
| “Old” target items | 0.75 (0.19) | 0.70 (0.16) | 0.53 (0.19) | 0.50 (0.20) |
| Easy lure items | 0.57 (0.17) | 0.66 (0.18) | 0.42 (0.19) | 0.48 (0.21) |
| Medium lure items | 0.54 (0.17) | 0.53 (0.17) | 0.42 (0.21) | 0.44 (0.21) |
| Hard lure items | 0.26 (0.15) | 0.39 (0.17) | 0.49 (0.17) | 0.44 (0.20) |

Mean (standard deviation) for accuracy and confidence of older adults (14 *APOE4* carriers, 43 *APOE4* non-carriers separately).

We then investigated the influence of recognition-demand level and the interaction of recognition-demand level by *APOE4* group on recognition confidence (see Table 5 for descriptive values, Figure 5b, and Supplementary Table S8). Here we again assessed a U-shaped trajectory additionally to the linear trajectory of confidence across demand levels. The model revealed that confidence decreased with increasing recognition-task demand in a linear (β=-0.36 [95% CI –0.45, –0.28], t=-8.25, *p*<0.001) manner, and additionally showed increase for hard levels, meaning a U-shaped manner (β=0.25 [95% CI 0.14, 0.35], t=4.65, *p*<0.001). There was no significant main effect of the AT-Term or *APOE4* group and no significant interaction with recognition-demand level (all *p*>0.05, see Supplementary Table S8 for statistics).

Additionally, we investigated the influence of accuracy and the interaction of recognition-demand level by *APOE4* group by accuracy on recognition confidence (see Table 5 for descriptive values, Figure S3 and S4, and Supplementary Table S9). The model revealed that confidence was higher for correct compared to incorrect responses (β=0.29 [95% CI 0.22, 0.36], t=7.70, *p*<0.001). There was an interaction effect of recognition demand by accuracy (β=-0.25 [95% CI –0.32, –0.17], t=-6.58, *p*<0.001, Cohen’s f²=0.15), with confidence for correct responses decreasing and confidence for incorrect responses increasing with higher demand level. Further, there was a three-way interaction of recognition-demand level by *APOE4* group by accuracy (β=-0.28 [95% CI –0.42, –0.30], t=-3.70, *p*<0.001, Cohen’s f²=0.02), with *APOE4* carriers compared to non-carriers showing a steeper increase in confidence across recognition-task demand levels for incorrect responses (estimate = –0.11 [95% CI –0.17, –0.04], *p*-FDR = .002) and descriptively a steeper decline in confidence across recognition-task demand levels for correct responses (estimate=0.06 [95% CI –0.02, 0.12], t=1.89, *p*-FDR=0.060).

VLMT recognition performance was not associated with *APOE4* group or the AT-Term (all *p*>0.05, see Supplementary Table S11 for statistics), RCFT recognition performance was not associated with *APOE4* group but with the Alzheimer’s pathology ratio, showing worse performance with more pathology (β=-0.47 [95% CI –0.76, –0.18], t=-3.30, *p*=0.002, see Supplementary Table S12 for statistics).

Further, we investigated the effect of recall-demand level and the interaction of recall-demand by *APOE4* group, and the effect of the AT-Term on recall performance (see Figure 5c and Supplementary Table S12), which was based on a recall scoring system (see Supplementary Methods S1 and Table S1). A mean recall score of cued and free recall was used (correlation of cued and free recall: r = 0.71 [95% CI 0.55, 0.82], t=7.41, *p*<0.001). The model revealed that performance decreased with increasing recall-task demand (β=-0.29 [95% CI –0.38, –0.21], t=-6.73, *p*<0.001). There was no significant main effect of the AT-Term or *APOE4* group and no significant interaction with recognition demand (all *p*>0.05, see Supplementary Table S12 for statistics).

Additionally, we investigated the influence of *APOE4* group and the AT-Term on recall error (see Figure 4d and Supplementary Table S13). There was no significant main effect of the AT-Term or *APOE4* group (all *p*>0.05, see Supplementary Table S13 for statistics).

Taken together, there were no main effects of *APOE4* carriership or Alzheimer’s pathology in older adults regarding recognition accuracy and confidence, recall score or recall error. In *APOE4* carriers, however, accuracy declined more steeply than in non-carriers with increasing recognition-task demand. *APOE4* carriers further showed a steeper increase in confidence across recognition-task demand levels for incorrect responses compared to non-carriers.

Third, only in older adults, we conducted correlation analysis between EMCOMP recognition accuracy and recall score with established episodic memory scores of the VLMT, RCFT, FCSRT, and WMS, as well as with the SDMT for processing speed, the RWT for executive functioning, and the CERAD-Plus for general cognition. We used false-discovery-rate corrected *p*-values to account for multiple comparisons.

For recognition accuracy, EMCOMP corrected hit rate was significantly correlated with VLMT corrected hit rate, which, however, did not survive FDR-correction (r =0.26 [95% CI 0.00, 0.49], t=1.99, *p*_FDR_=0.102), but here was no significant correlation with RCFT corrected hit rate (r =-0.10 [95% CI –0.35, 0.17 t=-0.72, *p*_FDR_=0.478). Mean EMCOMP recall score was significantly positively correlated with all four established episodic memory recall scores (see Figure 6 a – d), namely VLMT (r =0.58 [95% CI 0.37, 0.73 t=5.22, *p*_FDR_<0.001), FCSRT (r =0.35 [95% CI 0.10, 0.56 t=2.79, *p*_FDR_=0.007), RCFT (r =0.38 [95% CI 0.13, 0.58 t=3.02, *p*_FDR_=0.005), and WMS (r =0.44 [95% CI 0.20, 0.63 t=3.60, *p*_FDR_=0.001).

**Figure 6.**
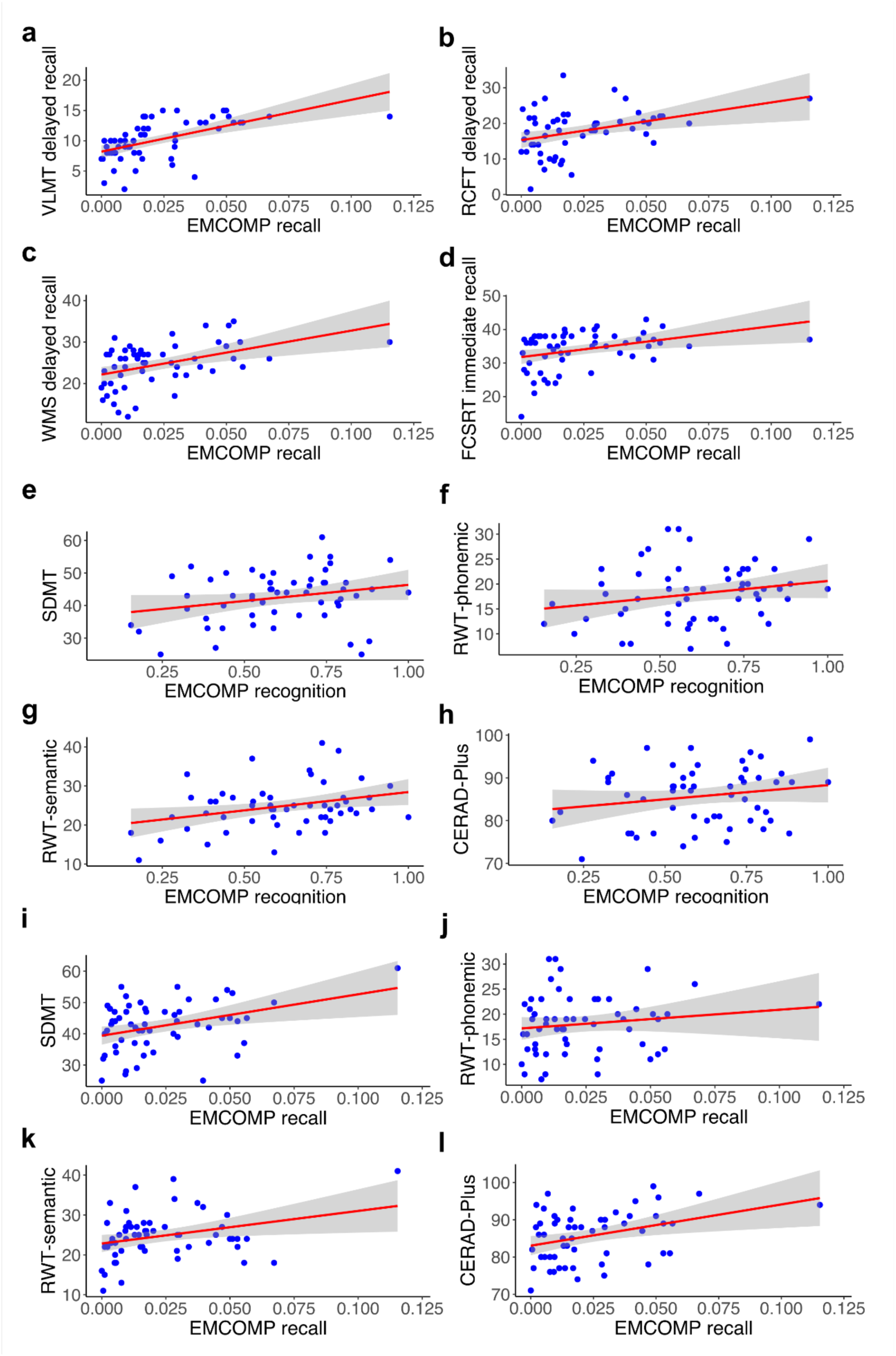
Correlations of EMCOMP performance and established cognitive tests. For recall, analyses were repeated when excluding a recall score > 0.12. The results remained significant, and the value is not categorized as outlier, as it is within 2 standard deviations, which was the criterion for outliers. a) EMCOMP recall performance and VLMT recall performance. b) EMCOMP recall performance and RCFT recall performance. c) EMCOMP recall performance and WMS recall performance. d) EMCOMP recall performance and FCSRT recall performance. e) EMCOMP recognition performance (corrected hit rate) and SDMT performance. f) EMCOMP recognition performance (corrected hit rate) and RWT-phonemic performance. g) EMCOMP recognition performance (corrected hit rate) and RWT-semantic performance. h) EMCOMP recognition (corrected hit rate) performance and CERAD-Plus performance. i) EMCOMP recall performance and SDMT performance. j) EMCOMP recall performance and RWT-phonemic performance. k) EMCOMP recall performance and RWT-semantic performance. l) EMCOMP recall performance and CERAD-Plus performance.

Regarding further cognitive functions, EMCOMP corrected hit rate was not significantly correlated with SDMT, RWT phonemic or semantic fluency, or CERAD-Plus (see Table 6 for statistics and Figure 6 e – h). Mean recall score was significantly positively correlated with SDMT, RWT semantic fluency, and CERAD-Plus, but not RWT phonemic fluency (see Table 6 for statistics and Figure 6 i – l).

**Table 6.** Correlation of EMCOMP recognition and recall score with cognitive tests.

| EMCOMP | Cognitive test | r | 95% CI | t | $p_{FDR}$ |
| --- | --- | --- | --- | --- | --- |
| recognition | SDMT | 0.25 | -0.02, 0.48 | 1.88 | 0.132 |
| recognition | RWT phonemic fluency | 0.22 | -0.04, 0.45 | 1.67 | 0.133 |
| recognition | RWT semantic fluency | 0.32 | 0.06, 0.53 | 2.49 | 0.063 |
| recognition | CERAD-Plus | 0.20 | -0.08, 0.44 | 1.44 | 0.156 |
| recall score | SDMT | 0.37 | 0.12, 0.57 | 2.91 | 0.015 |
| recall score | RWT phonemic fluency | 0.14 | -0.13, 0.39 | 1.04 | 0.301 |
| recall score | RWT semantic fluency | 0.31 | 0.05, 0.53 | 2.40 | 0.027 |
| recall score | CERAD-Plus | 0.36 | 0.10, 0.57 | 2.79 | 0.015 |

Correlation $r$ and the corresponding 95% confidence interval (CI), $t$ -value, and false-discovery-rate (FDR) corrected $p$ -value.

## 4. Discussion

### 4.1. Summary

Our paradigm incorporated multiple levels of task demand by manipulating the specificity of episodic memory retrieval. This approach allowed us to detect early subtle cognitive deficits and to provide novel empirical insights that complement theories of diminished neural resources with age and Alzheimer’s disease risk. Young adults showed higher recognition accuracy and confidence, a higher recall score, and a lower recall error rate compared to older adults. Importantly, we found a steeper decline in recognition accuracy and a steeper increase in high-confidence errors during recognition in older compared to young adults and in older *APOE4* carriers compared to non-carriers with higher recognition-task demand. There were, however, no associations with early plasma-based Alzheimer’s pathology. Lastly, EMCOMP performance was positively correlated with established cognitive tests.

### 4.2. Retrieval performance in young and older adults

Our first aim was to assess retrieval performance across multiple task-demand levels for young and older cognitively unimpaired adults. In older adults, the recall score declined from gist to detail and recognition accuracy decreased from novel to “old” target items to easy, medium, and hard lure conditions. Young adults showed the same pattern, except that easy lures were rejected more accurately than completely novel items. Our results align with prior studies in young and older adults regarding recall^75,76^ and delayed recognition^57,58^ with different retrieval-demand levels. The increasingly worse performance in the recognition task could be related to the increasing cognitive challenge caused by similarity between “old” target items and lures, as item recognition relies on familiarity and gist, whereas lure discrimination depends on recollection of detailed information^77–79^. The present study extends prior work by a more fine-grained demand manipulation via similarity using three lure levels, offering five recognition-demand levels in total. Additionally, using sentence-based stimuli creates a novel episodic retrieval paradigm for broader future application in memory research.

As expected from previous aging studies^80–84^, young adults showed higher recognition and recall performance than older adults. Older adults already performed around chance level for medium lures, whereas young adults only did so for hard lures. Older adults even performed considerably below chance level for hard lures. Prior studies reported an age-related decline in the ability of mnemonic discrimination and memory specificity^58,85,86^. This decline might contribute to a higher cognitive challenge for older adults in lure discrimination, where the identification of differences in specific details is required. Interestingly, older adults showed a steeper decline in recognition accuracy as task difficulty increased, possibly reflecting reduced ability to cope with increasing cognitive demands, in accordance with the proposed model by Reuter-Lorenz & Cappell^39^. Our findings are further consistent with results of a metaanalysis where a larger age difference for recollection-based recognition (discrimination of related and similar items) than for familiarity-based recognition was reported^87^.

The results of the current study are also consistent with reports suggesting that with age, gist retrieval in recognition tasks is preserved, while the retrieval of specific detail, needed for lure discrimination, declines^88–91^. For example, Abadie and colleagues conducted a study in which young and older adults studied a list of words which were related and unrelated, representing strong and weak gist relations, respectively. Both age groups successfully retrieved the gist but older adults particularly struggled in retrieving specific detail causing increased recognition errors^88^. Interestingly, a steeper decline in recognition accuracy with older age was not only reported for increasing similarity between items in visual tasks^88^, but also with increasing semantic ambiguity and noise for auditory tasks^10^, suggesting a modality-independent effect of increasing task demand.

Recognition confidence ratings reflected findings from prior literature^83,92,93^. Interestingly, while confidence generally decreased with increasing recognition-task demand, older adults, but not young adults, showed an increase in confidence at the highest difficulty levels, where they gave most incorrect responses. These findings align with the results from a study by Shing and colleagues^83^ on high-confidence memory errors in an associative recognition paradigm. Even though older adults performed comparably to children when recognising a word pair, they rated their confidence in errors higher than children did. Age-related differences in memory confidence, particularly the increase of high-confidence errors in older adults, could be attributed to difficulties in binding different components of an episode into a holistic, coherent memory representation^93^. Due to reduced memory specificity, older adults rely more on the gist of an episode when making memory judgments. Under high recognition demands with high item similarity, this reliance becomes problematic, as difficulties in retrieving specific details hinder lure discrimination and increase the likelihood of high-confidence errors^92^.

Surprisingly, young adults showed a steeper decline in recall performance across increasing retrieval demands (from gist to detail) compared to older adults. However, this apparent interaction effect should be interpreted with caution, as older adults’ performance approached floor levels at both recall-demand levels. Because their gist-recall performance was already low, further decline at the detail level could not be reliably detected. The paradigm, requiring the verbal recall of numerous information-rich sentences, may have exceeded older adults’ processing capacity. When task demands surpass available resources, cognitive effort decreases and disengagement occurs^39,40^. Thus, older adults may reach cognitive saturation earlier due to reduced neural resources. These assumptions warrant verification through neuroimaging studies.

### 4.3. Retrieval performance in older adults related to *APOE4* genotype and Alzheimer’s pathology

Our second aim was to assess the influence of the *APOE4* genotype and plasma-based Alzheimer’s pathology burden on retrieval performance in older adults. We found an effect of *APOE4* on recognition, but not recall performance. Specifically, *APOE4* carriers showed a steeper decline in recognition accuracy and a steeper increase in confidence across recognition-task demand levels for incorrect responses with higher recognition-task demand. In contrast, no *APOE4*-related effects emerged in the VLMT or RCFT recognition measures, suggesting that the EMCOMP task with multiple levels of task demand may be more sensitive to subtle, early deficits in retrieval processes.

A study by Rajah and colleagues^50^ employed an intentional spatial context encoding paradigm in middle-aged adults with a family history of sporadic Alzheimer’s disease, the *APOE4* allele, and controls. There were two demand levels, as participants either encoded six items (easy) or twelve items (hard). The study found no differences in retrieval accuracy between groups, but differences in activation between groups and demand levels in the hippocampus and parietal cortex. However, there were no interaction effects of group by demand level on activation. Interestingly, participants with Alzheimer’s risk factors exhibited more hippocampal activation during easy encoding trials compared to controls, and higher hippocampal activity during encoding was correlated with better subsequent memory performance. The question remains, whether introducing additional levels of task demand, as in our study, would have revealed interaction effects that were not detectable with only two demand levels. A further study^94^ in older adults with mild cognitive impairment reported higher functional connectivity between the hippocampus and parietal cortex in *APOE4* carriers compared to non-carriers, which was positively associated with a memory composite score in the carrier group. While this could hint towards functional compensation, other studies reported the opposite findings^95^. Overall, the role of *APOE* in aging and Alzheimer’s disease is multifaceted and remains incompletely understood^96^, but large cohort studies underline that memory decline is a key feature among cognitively unimpaired *APOE4* carriers^97^ and thus underscore the potential of tasks with varying demand levels for identifying nuanced tipping-points in performance changes.

There were no associations between plasma biomarkers related to Alzheimer’s disease (AT-Term = 1/ (Aβ_1-42_/Aβ_1-40_) * p-tau_217_) and EMCOMP recognition or recall performance, nor with VLMT recognition. However, in the RCFT recognition task, a higher plasma-based AT-Term was associated with worse performance. On the one hand, the degree of pathology in this cognitively healthy sample may not yet be sufficient to impact performance or the blood-based markers may not be sensitive enough in this healthy cohort. On the other hand, the absence of behavioral differences despite variability in the AT-Term could be related to functional compensation – that is, comparable behavioral performance despite differences in pathology burden, which could be compensated via specific neural activation or connectivity patterns^39,49^. In a previous study^98^ for instance, the authors used a letter Sternberg working memory paradigm with three demand levels (one, three, or six letters at the same time). There were no differences in accuracy between Aβ-positive and Aβ-negative cognitively unimpaired older adults, but Aβ-positive individuals showed stronger parametric brain activity increases with higher demand level during the delay period in frontoparietal regions, which was interpreted as compensatory upregulation and additional activation. Another study^99^ reported a parametric increase in brain activity in Aβ-positive cognitively unimpaired older adults during encoding that was positively correlated with episodic memory task performance, specifically the recall of detail information. Recent findings using multivariate bayesian decoding in a lifespan sample suggest that age increased the likelihood that higher activation patterns in the cuneal region during problem solving provided non-redundant information^100^. If higher activation reflects functional compensation as a short-term employment of additional neural resources to meet task demands or if it is part of a pathological cascade of hyperactivation and loss of modulation remains an open question^45^.

Overall, the observed interactions of *APOE4* status and recognition-task demand on performance may reflect early genotype-related neural resource limitations or alterations in resource allocation via, e.g., decreased network efficiency that precede effects of early pathology with age^101–104^. Future neuroimaging studies employing the EMCOMP paradigm are needed to address this question.

### 4.4. Validation of the retrieval task

Our third aim was to assess the relation of the performance in the EMCOMP task to performance in established cognitive tests in older adults. Regarding recognition performance, no significant associations were found between the corrected hit rates for EMCOMP and the widely-used memory measures VLMT and RCFT. This could likely reflect, on the one hand, the higher difficulty of the EMCOMP task with additional lure items and, on the other hand, ceiling effects observed in the traditional tests with fewer items and without lures.

For recall, EMCOMP performance showed positive correlations with the established memory recall measures VLMT, FCSRT, RCFT, and WMS, indicating convergent validity for retrieval processes. Beyond memory-specific tasks, recall but not recognition performance was positively associated with the SDMT, RWT semantic fluency, and the CERAD-Plus composite score. Further, there were positive associations between EMCOMP measures and the covariate working memory capacity, as reported in the supplementary tables. These broader associations suggest that successful retrieval in EMCOMP may rely on executive and semantic processes in addition to episodic memory, consistent with the task’s higher cognitive demands.

### 4.5. Limitations

Several limitations of the present study should be noted. First, our cohort was relatively homogeneous in demographic and educational background, which may limit the generalizability of the findings. Second, the sentences used in the EMCOMP task were not fully naturalistic, potentially constraining ecological validity, meaning the generalizability to real-life settings, and are restricted to the German language. Using our method of item construction, we were, however, able to incorporate more social, emotional, perceptual, spatial, and temporal details. Further, we assumed that increasing sentence similarity at recognition would lead to a parametric increase in task demand, although this cannot be demonstrated directly within the current design. Third, recall performance in older adults showed evidence of floor effects, indicating that this condition may have been too demanding for this age group and possibly limiting the interpretability of group differences. Fourth, while the current study focused on behavioral performance, no neuroimaging data were yet available. As a next step, we plan to implement the EMCOMP task in the MRI scanner to examine potential functional compensation mechanisms^36,39^, that is, preserved behavioral performance despite Alzheimer’s risk or pathology linked to altered activation or connectivity patterns. Additionally, the inclusion of tau-PET imaging, particularly targeting the medial temporal lobe, will allow a closer investigation of the neural correlates of episodic memory integrity and very early Alzheimer’s pathology^51^. Finally, future studies should account for individual differences in cognitive reserve proxies, additionally to functional compensation during the task, which may modulate the relation between pathology, neural activation or connectivity, and behavioral performance.

## 5. Conclusion

Our findings indicate that age– and *APOE4*-related differences in episodic memory retrieval with increasing task demands could be driven by the rising cognitive challenge associated with higher levels of specificity. Vulnerable groups, meaning older adults and *APOE4* carriers, appear less able to overcome these increasing demands, potentially reflecting reduced neural resources. Importantly, the highest demand levels proved too difficult for older adults; as a result, the EMCOMP task effectively spans the full range from manageable to unmanageable challenge. This underscores its suitability for investigating functional compensation with neuroimaging, offering a sensitive testbed for theories of compensatory mechanisms in preclinical stages of cognitive decline.

## Declarations

### Ethics approval and consent to participate

All study procedures and experimental protocols were approved by the Ethics Committee of the Medical Faculty, Otto-von-Guericke University Magdeburg (200/19) and were carried out in accordance with the Declaration of Helsinki. All participants provided written informed consent.

### Consent for publication

Not applicable.

### Availability of data and materials

The datasets generated and/or analysed during the current study are not publicly available due to the inclusion of sensitive participant information and privacy concerns but are available from the corresponding author on reasonable request for replication. Code for statistical analysis is available at https://github.com/fislarissa/EMCOMP_demand.

### Authors’ contributions

Conceptualisation: LF, JP, RO, AW, AM. Methodology: LF, JP, AM. Formal analysis: LF, FA. Data Acquisition: FA, LF, BSW, NB. Data Processing: LF, FA, BSW, NB, GB, BM, HE. Visualisation: LF. Supervision: LF, JP, AM. Project administration: LF, BSW, ED, AM. Funding acquisition: ED, AM. Resources: ED, AM. Writing original draft: LF, FA, AM. Writing – review and editing: All authors. All authors read and approved the final manuscript.

## Data Availability

All data produced in the present study are available upon reasonable request to the authors.

https://doi.org/10.17605/OSF.IO/E4MC9

https://github.com/fislarissa/EMCOMP_demand

## List of abbreviations

AD: Alzheimer’s Disease
Aβ: Amyloid-beta
*APOE4*: Apolipoprotein-E4
CI: Confidence Interval
DZNE: German Center for Neurodegenerative Diseases
EMCOMP: Episodic Memory & COMPensation study

## Acknowledgements

We want to thank the participants for their time and effort, Julie Piechnik and Julia Pontow for their help, and the RTG 2413 SynAGE for supporting Larissa Fischer’s research stay in Boston, USA.

## References

[1] McKhann GM, Knopman DS, Chertkow H, Hyman BT, Jack CR Jr, Kawas CH, et al. The diagnosis of dementia due to Alzheimer’s disease: recommendations from the National Institute on Aging-Alzheimer’s Association workgroups on diagnostic guidelines for Alzheimer’s disease. Alzheimers Dement. 2011;7(3):263–269.

[2] Salthouse TA. Trajectories of normal cognitive aging. Psychol Aging. 2019;34(1):17–24.

[3] Grady C. The cognitive neuroscience of ageing. Nature Reviews Neuroscience. 2012;13(7):491–505.

[4] Luo L, Craik FIM. Aging and Memory: A Cognitive Approach. The Canadian Journal of Psychiatry. Published online 2008. doi:10.1177/070674370805300603

[5] Babcock RL, Salthouse TA. Effects of increased processing demands on age differences in working memory. Psychol Aging. 1990;5(3):421–428.

[6] Wang M, Gamo NJ, Yang Y, Jin LE, Wang XJ, Laubach M, et al. Neuronal Basis of Age-Related Working Memory Decline. Nature. 2011;476(7359):210.

[7] Davis SW, Zhuang J, Wright P, Tyler LK. Age-related sensitivity to task-related modulation of language-processing networks. Neuropsychologia. 2014;63:107–115.

[8] Salthouse TA, Pink JE. Why is working memory related to fluid intelligence? Psychonomic Bulletin & Review. 2013;15(2):364–371.

[9] Loaiza VM, Oftinger AL, Camos V. How does Working Memory Promote Traces in Episodic Memory? Journal of Cognition. 2023;6(1). doi:10.5334/joc.245

[10] Koeritzer MA, Rogers CS, Van Engen KJ, Peelle JE. The Impact of Age, Background Noise, Semantic Ambiguity, and Hearing Loss on Recognition Memory for Spoken Sentences. Published online March 15, 2018. doi:10.1044/2017_JSLHR-H-17-0077

[11] López-Otín C, Blasco MA, Partridge L, Serrano M, Kroemer G. Hallmarks of aging: An expanding universe. Cell. 2023;186(2):243–278.

[12] Bergado JA, Almaguer W. Aging and Synaptic Plasticity: A Review. Neural Plasticity. 2002;9(4):217–232.

[13] Jansen WJ, Ossenkoppele R, Knol DL, Tijms BM, Scheltens P, Verhey FRJ, et al. Prevalence of cerebral amyloid pathology in persons without dementia: A meta-analysis. JAMA. 2015;313(19):1924.

[14] Moscoso A, Heeman F, Raghavan S, Costoya-Sánchez A, van Essen M, Mainta I, et al. Frequency and Clinical Outcomes Associated With Tau Positron Emission Tomography Positivity. JAMA. 2025;334(3):229–242.

[15] Braak H, Braak E. Neuropathological stageing of Alzheimer-related changes. Acta Neuropathol. 1991;82(4):239–259.

[16] Braak H, Alafuzoff I, Arzberger T, Kretzschmar H, Del Tredici K. Staging of Alzheimer disease-associated neurofibrillary pathology using paraffin sections and immunocytochemistry. Acta Neuropathol. 2006;112(4):389–404.

[17] Villeneuve S, Rabinovici GD, Cohn-Sheehy BI, Madison C, Ayakta N, Ghosh PM, et al. Existing Pittsburgh Compound-B positron emission tomography thresholds are too high: statistical and pathological evaluation. Brain. 2015;138(Pt 7):2020–2033.

[18] Thal DR, Rüb U, Orantes M, Braak H. Phases of A beta-deposition in the human brain and its relevance for the development of AD. Neurology. 2002;58(12):1791–1800.

[19] Ossenkoppele R, Pichet Binette A, Groot C, Smith R, Strandberg O, Palmqvist S, et al. Amyloid and tau PET-positive cognitively unimpaired individuals are at high risk for future cognitive decline. Nat Med. 2022;28(11):2381–2387.

[20] Mayeux R. Epidemiology of neurodegeneration. Annu Rev Neurosci. 2003;26:81–104.

[21] Liu CC, Liu CC, Kanekiyo T, Xu H, Bu G. Apolipoprotein E and Alzheimer disease: risk, mechanisms and therapy. Nat Rev Neurol. 2013;9(2):106–118.

[22] Villemagne VL, Rowe CC. Long night’s journey into the day: amyloid-β imaging in Alzheimer’s disease. J Alzheimers Dis. 2013;33 Suppl 1:S349–S359.

[23] Selkoe DJ, Hardy J. The amyloid hypothesis of Alzheimer’s disease at 25 years. EMBO Mol Med. 2016;8(6):595–608.

[24] Lancaster C, Berens S, Daly J, Rusted J, Bird CM. Perceptual discrimination of complex objects: Apolipoprotein E e4 gene-dose effects in mid-life. Alzheimer’s & Dementia. 2025;21(6):e70246.

[25] Coughlan G, Coutrot A, Khondoker M, Minihane AM, Spiers H, Hornberger M. Toward personalized cognitive diagnostics of at-genetic-risk Alzheimer’s disease. Proceedings of the National Academy of Sciences. 2019;116(19):9285–9292.

[26] Lancaster C, Tabet N, Rusted J. The Elusive Nature of APOE ε4 in Mid-adulthood: Understanding the Cognitive Profile. Journal of the International Neuropsychological Society: JINS. 2017;23(3). doi:10.1017/S1355617716000990

[27] Lissaman R, Anjum S, Quaiattini A, Rajah MN. APOE ε4-related differences in brain structure, function, and connectivity at midlife: A scoping review. J Prev Alzheimers Dis. 2025;12(9):100364.

[28] Sinha N, Berg CN, Tustison NJ, Shaw A, Hill D, Yassa MA, et al. APOE ε4 status in healthy older African Americans is associated with deficits in pattern separation and hippocampal hyperactivation. Neurobiol Aging. 2018;69:221–229.

[29] Caselli RJ, Dueck AC, Osborne D, Sabbagh MN, Connor DJ, Ahern GL, et al. Longitudinal Modeling of Age-Related Memory Decline and the APOE ε4 Effect. Published online July 16, 2009. doi:10.1056/NEJMoa0809437

[30] Gellersen HM, Coughlan G, Hornberger M, Simons JS. Memory precision of object-location binding is unimpaired in APOE ε4-carriers with spatial navigation deficits. Brain Commun. 2021;3(2):fcab087.

[31] Morrison C, Oliver MD, Berry V, Kamal F, Dadar M. The influence of APOE status on rate of cognitive decline. GeroScience. 2024;46(3):3263–3274.

[32] Lu M, Kim MJ, Collins EC, Shcherbinin S, Ellinwood AK, Yokoi Y, et al. Posttreatment Amyloid Levels and Clinical Outcomes Following Donanemab for Early Symptomatic Alzheimer Disease: A Secondary Analysis of the TRAILBLAZER-ALZ 2 Randomized Clinical Trial. JAMA Neurol. Published online October 13, 2025. doi:10.1001/jamaneurol.2025.3869

[33] Sims JR, Zimmer JA, Evans CD, Lu M, Ardayfio P, Sparks J, et al. Donanemab in Early Symptomatic Alzheimer Disease: The TRAILBLAZER-ALZ 2 Randomized Clinical Trial. JAMA. 2023;330(6):512–527.

[34] Mintun MA, Lo AC, Duggan Evans C, Wessels AM, Ardayfio PA, Andersen SW, et al. Donanemab in Early Alzheimer’s Disease. N Engl J Med. 2021;384(18):1691–1704.

[35] Stern Y, Albert M, Barnes CA, Cabeza R, Pascual-Leone A, Rapp PR. A framework for concepts of reserve and resilience in aging. Neurobiol Aging. 2023;124:100–103.

[36] Cabeza R, Albert M, Belleville S, Craik FIM, Duarte A, Grady CL, et al. Maintenance, reserve and compensation: the cognitive neuroscience of healthy ageing. Nat Rev Neurosci. 2018;19(11):701–710.

[37] Reuter-Lorenz PA, Park DC. How does it STAC up? Revisiting the scaffolding theory of aging and cognition. Neuropsychol Rev. 2014;24(3):355–370.

[38] Park DC, Reuter-Lorenz P. The adaptive brain: aging and neurocognitive scaffolding. Annu Rev Psychol. 2009;60:173–196.

[39] Reuter-Lorenz PA, Cappell KA. Neurocognitive Aging and the Compensation Hypothesis. Curr Dir Psychol Sci. 2008;17(3):177–182.

[40] Peelle JE. Listening effort: How the cognitive consequences of acoustic challenge are reflected in brain and behavior. Ear Hear. 2018;39(2):204–214.

[41] Silvestrini N, Musslick S, Berry AS, Vassena E. An integrative effort: Bridging motivational intensity theory and recent neurocomputational and neuronal models of effort and control allocation. Psychol Rev. 2023;130(4):1081–1103.

[42] Kennedy KM, Rodrigue KM, Bischof GN, Hebrank AC, Reuter-Lorenz PA, Park DC. Age trajectories of functional activation under conditions of low and high processing demands: an adult lifespan fMRI study of the aging brain. Neuroimage. 2015;104:21–34.

[43] Aschenbrenner AJ, Crawford JL, Peelle JE, Fagan AM, Benzinger TLS, Morris JC, et al. Increased cognitive effort costs in healthy aging and preclinical Alzheimer’s disease. Psychol Aging. 2023;38(5):428–442.

[44] Park DC, Polk TA, Hebrank AC, Jenkins LJ. Age differences in default mode activity on easy and difficult spatial judgment tasks. Front Hum Neurosci. 2010;3:75.

[45] Corriveau-Lecavalier N, Adams JN, Fischer L, Molloy EN, Maass A. Cerebral hyperactivation across the Alzheimer’s disease pathological cascade. Brain Commun. 2024;6(6):fcae376.

[46] Peelle JE, Troiani V, Wingfield A, Grossman M. Neural processing during older adults’ comprehension of spoken sentences: age differences in resource allocation and connectivity. Cereb Cortex. 2010;20(4):773–782.

[47] Piquado T, Isaacowitz D, Wingfield A. Pupillometry as a measure of cognitive effort in younger and older adults. Psychophysiology. 2010;47(3):560–569.

[48] DeCaro R, Peelle JE, Grossman M, Wingfield A. The Two Sides of Sensory–Cognitive Interactions: Effects of Age, Hearing Acuity, and Working Memory Span on Sentence Comprehension. Front Psychol. 2016;7:167409.

[49] Haitas N, Dubuc J, Massé-Leblanc C, Chamberland V, Amiri M, Glatard T, et al. Registered report: Age-preserved semantic memory and the CRUNCH effect manifested as differential semantic control networks: An fMRI study. PLOS ONE. 2024;19(6):e0289384.

[50] Rajah MN, Wallace LMK, Ankudowich E, Yu EH, Swierkot A, Patel R, et al. Family history and APOE4 risk for Alzheimer’s disease impact the neural correlates of episodic memory by early midlife. Neuroimage Clin. 2017;14:760–774.

[51] Maass A, Lockhart SN, Harrison TM, Bell RK, Mellinger T, Swinnerton K, et al. Entorhinal Tau Pathology, Episodic Memory Decline, and Neurodegeneration in Aging. J Neurosci. 2018;38(3):530–543.

[52] Garo-Pascual M, Gaser C, Zhang L, Tohka J, Medina M, Strange BA. Brain structure and phenotypic profile of superagers compared with age-matched older adults: a longitudinal analysis from the Vallecas Project. Lancet Healthy Longev. 2023;4(8):e374–e385.

[53] Just MA, Carpenter PA, Keller TA, Eddy WF, Thulborn KR. Brain activation modulated by sentence comprehension. Science. 1996;274(5284):114–116.

[54] Fiebach CJ, Schlesewsky M, Lohmann G, von Cramon DY, Friederici AD. Revisiting the role of Broca’s area in sentence processing: syntactic integration versus syntactic working memory. Human brain mapping. 2005;24(2). doi:10.1002/hbm.20070

[55] Behrenbruch N, Schwarck S, Schumann-Werner B, Molloy EN, Garcia-Garcia B, Hochkeppler A, et al. A physically and mentally active lifestyle relates to younger brain and cognitive age. GeroScience. Published online July 7, 2025:1–21.

[56] Fischer L. Preregistration: Episodic memory retrieval across different demand levels related to aging and Alzheimer’s disease pathology – a behavioral study. OSF. Published 2024. https://osf.io/68dkj/metadata/osf. Accessed October 20, 2025

[57] Karlsson AE, Wehrspaun CC, Sander MC. Item recognition and lure discrimination in younger and older adults are supported by alpha/beta desynchronization. Neuropsychologia. 2020;148:107658.

[58] Ilyés A, Paulik B, Keresztes A. Discrimination of semantically similar verbal memory traces is affected in healthy aging. Sci Rep. 2024;14(1):17971.

[59] Oxford Languages and Google – English. Published May 20, 2020. https://languages.oup.com/google-dictionary-en/. Accessed October 17, 2025

[60] DWDS. DWDS − Der deutsche Wortschatz von 1600 bis heute. Published 2024. https://www.dwds.de/. Accessed December 12, 2024

[61] Peirce J, Gray JR, Simpson S, MacAskill M, Höchenberger R, Sogo H, et al. PsychoPy2: Experiments in behavior made easy. Behav Res Methods. 2019;51(1):195–203.

[62] Malyutina S, Laurinavichyute A, Terekhina M, Lapin Y. No evidence for strategic nature of age-related slowing in sentence processing. Psychol Aging. 2018;33(7):1045–1059.

[63] Lehrl S. Mehrfachwahl-Wortschatz-Intelligenztest: MWT-B [Manual zum MWT-B].; 2005.

[64] GitHub – colinquirk/PsychopyChangeDetection: Change Detection in Psychopy. GitHub. Published 2020. https://github.com/colinquirk/PsychopyChangeDetection. Accessed October 17, 2025

[65] Rensink RA. Change detection. Annu Rev Psychol. 2002;53:245–277.

[66] Pashler H. Familiarity and visual change detection. Percept Psychophys. 1988;44(4):369–378.

[67] Cowan N. The magical number 4 in short-term memory: a reconsideration of mental storage capacity. Behav Brain Sci. 2001;24(1):87–114; discussion 114-185.

[68] Helmstaedter C, Durwen HF. The Verbal Learning and Retention Test. A useful and differentiated tool in evaluating verbal memory performance. Schweizer Archiv fur Neurologie und Psychiatrie. 1990;141(1). https://pubmed.ncbi.nlm.nih.gov/1690447/. Accessed October 17, 2025

[69] Meyers JE, Meyers KR. Rey complex figure test under four different administration procedures. The Clinical Neuropsychologist. Published online November 8, 2007:63–67.

[70] Wechsler D. WMS-R: Wechsler Memory Scale--Revised: Manual.; 1987.

[71] Buschke H. Cued recall in amnesia. Journal of clinical neuropsychology. 1984;6(4). doi:10.1080/01688638408401233

[72] Smith A. Symbol Digit Modalities Test: Manual.; 2002.

[73] Aschenbrenner S, Tucha O, Lange KW. Regensburger Wortflüssigkeits-Test (RWT). Verlag für Psychologie: Hogrefe; 2000.

[74] Welsh KA, Butters N, Mohs RC, Beekly D, Edland S, Fillenbaum G, et al. The Consortium to Establish a Registry for Alzheimer’s Disease (CERAD). Part V. A normative study of the neuropsychological battery. Neurology. 1994;44(4). doi:10.1212/wnl.44.4.609

[75] Sacripante R, Logie RH, Baddeley A, Della Sala S. Forgetting rates of gist and peripheral episodic details in prose recall. Mem Cognit. 2023;51(1):71–86.

[76] Rhodes S, Greene NR, Naveh-Benjamin M. Age-related differences in recall and recognition: a meta-analysis. Psychon Bull Rev. 2019;26(5):1529–1547.

[77] Friedman D, de Chastelaine M, Nessler D, Malcolm B. Changes in familiarity and recollection across the lifespan: an ERP perspective. Brain Res. 2010;1310:124–141.

[78] Yonelinas AP. The nature of recollection and familiarity: A review of 30 years of research. J Mem Lang. 2002;46(3):441–517.

[79] Mandler G. Recognizing: The judgment of previous occurrence. Psychol Rev. 1980;87(3):252–271.

[80] Abichou K, La Corte V, Bellegarde A, Nicolas S, Piolino P. How rich are false memories in a naturalistic context in healthy aging? Memory. 2022;30(3):262–278.

[81] Schacter DL, Koutstaal W, Norman KA. False memories and aging. Trends Cogn Sci. 1997;1(6):229–236.

[82] McCabe DP, Roediger HL 3rd,McDaniel MA, Balota DA. Aging reduces veridical remembering but increases false remembering: neuropsychological test correlates of remember-know judgments. Neuropsychologia. 2009;47(11):2164–2173.

[83] Shing YL, Werkle-Bergner M, Li SC, Lindenberger U. Committing memory errors with high confidence: older adults do but children don’t. Memory. 2009;17(2):169–179.

[84] Cangelosi M, Rinaldi L, Dijkstra T, Palladino P, Cavallini E. Older adults produce more verbal false memories than younger adults: is it semantics or executive functioning? Aging Clinical and Experimental Research. 2025;37(1):1–11.

[85] Brock Kirwan C, Hartshorn A, Stark SM, Goodrich-Hunsaker NJ, Hopkins RO, Stark CEL. Pattern separation deficits following damage to the hippocampus. Neuropsychologia. 2012;50(10):2408–2414.

[86] Yassa MA, Lacy JW, Stark SM, Albert MS, Gallagher M, Stark CEL. Pattern separation deficits associated with increased hippocampal CA3 and dentate gyrus activity in nondemented older adults. Hippocampus. 2011;21(9):968–979.

[87] Fraundorf SH, Hourihan KL, Peters RA, Benjamin AS. Aging and recognition memory: A meta-analysis. Psychol Bull. 2019;145(4):339–371.

[88] Abadie M, Gavard E, Guillaume F. Verbatim and gist memory in aging. Psychol Aging. 2021;36(8):891–901.

[89] Brainerd CJ, Reyna VF. Gist is the grist: Fuzzy-trace theory and the new intuitionism. Dev Rev. 1990;10(1):3–47.

[90] Greene NR, Naveh-Benjamin M. A Specificity Principle of Memory: Evidence From Aging and Associative Memory. Psychol Sci. 2020;31(3):316–331.

[91] Grilli MD, Sheldon S. Autobiographical event memory and aging: older adults get the gist. Trends Cogn Sci. 2022;26(12):1079–1089.

[92] Greene NR, Naveh-Benjamin M. The time course of encoding specific and gist episodic memory representations among young and older adults. Journal of experimental psychology General. 2024;153(6). doi:10.1037/xge0001589

[93] Naveh-Benjamin M. Adult age differences in memory performance: tests of an associative deficit hypothesis. J Exp Psychol Learn Mem Cogn. 2000;26(5):1170–1187.

[94] De Marco M, Vallelunga A, Meneghello F, Varma S, Frangi AF, Venneri A. ApoE ε4 Allele Related Alterations in Hippocampal Connectivity in Early Alzheimer’s Disease Support Memory Performance. Curr Alzheimer Res. 2017;14(7):766–777.

[95] Bai F, Zhang Z, Watson DR, Yu H, Shi Y, Yuan Y, et al. Abnormal functional connectivity of hippocampus during episodic memory retrieval processing network in amnestic mild cognitive impairment. Biol Psychiatry. 2009;65(11):951–958.

[96] Jackson RJ, Hyman BT, Serrano-Pozo A. Multifaceted roles of APOE in Alzheimer disease. Nat Rev Neurol. 2024;20(8):457–474.

[97] Walters S, Contreras AG, Eissman JM, Mukherjee S, Lee ML, Choi SE, et al. Associations of Sex, Race, and Apolipoprotein E Alleles With Multiple Domains of Cognition Among Older Adults. JAMA Neurol. 2023;80(9):929–939.

[98] Oh H, Steffener J, Razlighi QR, Habeck C, Liu D, Gazes Y, et al. Aβ-related hyperactivation in frontoparietal control regions in cognitively normal elderly. Neurobiol Aging. 2015;36(12):3247–3254.

[99] Elman JA, Oh H, Madison CM, Baker SL, Vogel JW, Marks SM, et al. Neural compensation in older people with brain amyloid-β deposition. Nat Neurosci. 2014;17(10):1316–1318.

[100] Knights E, Henson RN, Morcom A, Mitchell DJ, Tsvetanov KA. Neural evidence of functional compensation for fluid intelligence in healthy ageing. Published online January 8, 2025. doi:10.7554/eLife.93327

[101] McDonough IM, Festini SB, Wood MM. Risk for Alzheimer’s disease: A review of long-term episodic memory encoding and retrieval fMRI studies. Ageing Res Rev. 2020;62:101133.

[102] Najm R, Jones EA, Huang Y. Apolipoprotein E4, inhibitory network dysfunction, and Alzheimer’s disease. Mol Neurodegener. 2019;14(1):24.

[103] Jones DT, Graff-Radford J, Lowe VJ, Wiste HJ, Gunter JL, Senjem ML, et al. Tau, amyloid, and cascading network failure across the Alzheimer’s disease spectrum. Cortex. 2017;97:143–159.

[104] Jones DT, Knopman DS, Gunter JL, Graff-Radford J, Vemuri P, Boeve BF, et al. Cascading network failure across the Alzheimer’s disease spectrum. Brain. 2016;139(Pt 2):547–562.

