## Supplementary Material for "Episodic memory retrieval with increasing task demand: Associations with age, *APOE4* genotype, and Alzheimer’s disease pathology"

[1] German Center for Neurodegenerative Diseases (DZNE), Magdeburg, Germany
[2] Institute of Cognitive Neurology and Dementia Research, Otto von Guericke University, Magdeburg, Germany
[3] Institute for Biology, Otto von Guericke University, Magdeburg, Germany
[4] Division of Nuclear Medicine, Department of Radiology & Nuclear Medicine, Faculty of Medicine, Otto von Guericke University, Magdeburg, Germany
[5] Department of Communication Sciences and Disorders and Department of Psychology, Northeastern University, Boston, USA
[6] Department of Psychology and Volen National Center for Complex Systems, Brandeis University, Waltham, USA
[7] Leibniz Institute for Neurobiology (LIN), Magdeburg, Germany
[8] Department of Psychiatry and Psychotherapy, University Medical Center, Georg-August University, Göttingen, Germany
[9] German Center for Neurodegenerative Diseases (DZNE), Göttingen, Germany

### Supplementary Methods S1. Recall scoring system

To evaluate the recall session, a scoring system was adapted based on prior research that distinguished between gist and detail information (Bird et al., 2015; Burke et al., 1992; Wessel et al., 2015). The scoring system separates the components of the sentences in two main categories: central (= gist sum score) and peripheral information (= detail sum score). The two scores represent the two recall demand levels. The gist sum score includes information about social relations (SOC - social central) and action (AC - action central) in the sentence. The detail sum score consists of five subcategories: social (SOP - social peripheral), emotional (EP - emotional peripheral), perceptual (PP - perceptual peripheral), spatial (SP - spatial peripheral) and temporal (TP - temporal peripheral) information. Each of the 75 encoded sentences was scored using the scoring system to predefine the maximum number of points possible for each category and the total sum scores, respectively. Then the free (90 minutes) and cued recall (100 minutes) were scored by giving one point for a correctly recalled piece of information in the respective category. Half a point was given when a word was recalled with similar content to the encoded (correct) word, e.g. saying "holiday" instead of "vacation". The total scores for each category were collected, as well as a total sum score for gist (total gist = SC + AC) and detail (total detail = SOP + EP + PP + SP + TP). The scoring also accounts for errors, including swaps and add ons. Swaps mean switching information from different encoded sentences; and add ons mean additional information that was not encoded. Two examples for a free recall scoring can be found in Table S1.

After scoring each recall type, the total scores were transformed into relative scores, meaning they were relative to the maximum possible score in the free or cued recall, respectively. Finally, a total recall score including information from both cued and free recall for either gist or detail information was built (free recall detail + cued recall detail; free recall gist cued recall gist). The relative total error score included swaps and add ons in cued and free recall in relation to the total recall score. Therefore, the relative error scores rather represent an error rate. If the error rate is greater than 1, it means the person recalled more information correctly than they made errors. If the rate equals 1, the person made as many errors as correct recalls and if the rate is smaller than 1 it indicates more errors than correct recalls.

**Table S1. Examples for the recall scoring**

| | | | | | | | | | |
|---|---|---|---|---|---|---|---|---|---|
| | **Example 1** | | | | | | | | |
| Example 1 | **Marvin** in **dunkelblauer Hose**, den sein **Vater** **unterstützt**, wird im **Sommerurlaub** **reich**. *Marvin in dark blue pants, whom his father supports, is getting rich on summer vacation.* | | | | | | | | |
| Scoring categories | **SOC** | **SOP** | **AC** | **EP** | **PP** | **SP** | **TP** | | |
| Maximum score | 2 | - | 1 | 1 | 2 | - | 1 | | |
| | **Free recall 1** | | | | | | | | |
| Recall example 1 | „**Jemand** wird **reich**…**unterstützt** von seinem **Vater** während der **Sommerferien**" *"Someone is getting rich… supported by his father during the summer holidays."* | | | | | | | | |
| Scoring categories | **SOC** | **SOP** | **AC** | **EP** | **PP** | **SP** | **TP** | **swaps** | **add on** |
| Scoring | 1,5 | - | 1 | 1 | 0 | - | 0,5 | - | - |
| | **Example 2** | | | | | | | | |
| Example 2 | **Oma Katharina** in ihrem **grünen** **Haus**, die ihre **Enkel** in den **Ferien** **verwöhnt**, ist **fürsorglich**. *"Grandma Katharina in her green house, who is pampering her grandchildren during the summer holidays, is caring."* | | | | | | | | |
| Scoring categories | **SOC** | **SOP** | **AC** | **EP** | **PP** | **SP** | **TP** | | |
| Maximum score | 2 | - | 1 | 1 | 2 | - | 1 | | |
| | **Free recall 2** | | | | | | | | |
| Recall example 2 | „**Oma Katharina** hat ihre **Enkel** bei den Hausaufgaben **unterstützt…im** **grünen** **Haus**" *"Grandma Katharina supported her grandchildren with their homework…in a green garden house."* | | | | | | | | |
| Scoring categories | **SOC** | **SOP** | **AC** | **EP** | **PP** | **SP** | **TP** | **swaps** | **add on** |
| Scoring | 2 | 1 | 0 | 0 | 1 | 1 | 0 | 1 | 1 |

Demonstration of scoring two sentences with the recall scoring system, showing the maximum scores possible and an example scoring for a free recall response in German with a literal translation in English below. The scoring system accounts for correctly recalled gist (central) information, details (peripheral) and recall errors (swaps and add ons). Abbreviations: SOC…social central, SOP…social peripheral, AC…action central, EP…emotional peripheral, PP…perceptual peripheral, SP…spatial peripheral, TP…temporal peripheral.

**Table S2. Linear model of effects of recognition-demand level on accuracy in young and older adults**

| | Accuracy | | | | | | | |
|---|---|---|---|---|---|---|---|---|
| *Predictors* | *Estimates* | *std. Error* | *std. Beta* | *standardized std. Error* | *CI* | *standardized CI* | *Statistic* | *p* |
| (Intercept) | 0.73 | 0.10 | 0.27 | 0.07 | 0.54 – 0.92 | 0.13 – 0.41 | 7.51 | **<0.001** |
| Recognition-demand level | -0.15 | 0.01 | -0.63 | 0.05 | -0.17 – -0.13 | -0.73 – -0.53 | -12.71 | **<0.001** |
| Age group [Older adults] | -0.08 | 0.02 | -0.35 | 0.09 | -0.12 – -0.04 | -0.52 – -0.18 | -3.99 | **<0.001** |
| Sex [Male] | -0.03 | 0.01 | -0.12 | 0.06 | -0.06 – 0.00 | -0.24 – 0.01 | -1.86 | 0.063 |
| Vocab score | -0.00 | 0.00 | -0.00 | 0.04 | -0.00 – 0.00 | -0.08 – 0.08 | -0.04 | 0.970 |
| Working memory score | 0.03 | 0.01 | 0.13 | 0.03 | 0.02 – 0.05 | 0.06 – 0.19 | 3.87 | **<0.001** |
| Recognition-demand level × Age group [Older adults] | -0.03 | 0.01 | -0.13 | 0.06 | -0.06 – -0.00 | -0.25 – -0.00 | -2.01 | **0.045** |
| **Random Effects** | | | | | | | | |
| $\sigma^2$ | 0.02 | | | | | | | |
| $\tau_{00\ participant}$ | 0.00 | | | | | | | |
| $N_{participant}$ | 92 | | | | | | | |
| Observations | 460 | | | | | | | |
| Marginal $R^2$ / Conditional $R^2$ | 0.567 / NA | | | | | | | |

Exploratory post-hoc analysis of the interaction effect with recognition-demand level as a factor revealed a significant difference between age groups for the third recognition-demand level (easy lures) ($t(434) = 5.179$, $p < 0.001$, SE = 0.035) and the fifth recognition-demand level (hard lures) ($t(224) = 3.242$, $p = 0.001$, SE = 0.035) using emmeans.

**Table S3. Linear model of effects of recognition-demand level on confidence in young and older adults**

| | Confidence | | | | | | | |
|---|---|---|---|---|---|---|---|---|
| *Predictors* | *Estimates* | *std. Error* | *std. Beta* | *standardized std. Error* | *CI* | *standardized CI* | *Statistic* | *p* |
| (Intercept) | 0.44 | 0.20 | 0.23 | 0.17 | 0.03 – 0.84 | -0.11 – 0.57 | 2.15 | **0.034** |
| Recognition-demand level | -0.07 | 0.01 | -0.32 | 0.05 | -0.09 – -0.05 | -0.42 – -0.22 | -6.30 | **<0.001** |
| Age group [Older adults] | -0.09 | 0.05 | -0.43 | 0.21 | -0.19 – -0.00 | -0.84 – -0.01 | -2.03 | **0.044** |
| Recognition-demand level U-shape | 0.01 | 0.01 | 0.04 | 0.06 | -0.02 – 0.04 | -0.07 – 0.16 | 0.74 | 0.460 |
| Sex [Male] | -0.07 | 0.03 | -0.33 | 0.14 | -0.13 – -0.01 | -0.61 – -0.06 | -2.39 | **0.019** |
| Vocab score | 0.00 | 0.00 | 0.06 | 0.09 | -0.00 – 0.01 | -0.13 – 0.24 | 0.63 | 0.532 |
| Working memory score | 0.03 | 0.02 | 0.13 | 0.07 | -0.00 – 0.06 | -0.02 – 0.28 | 1.74 | 0.086 |
| Recognition-demand level × Age group [Older adults] | -0.01 | 0.01 | -0.04 | 0.06 | -0.04 – 0.02 | -0.17 – 0.09 | -0.61 | 0.540 |
| Recognition-demand level U-shape × Age group [Older adults] | 0.05 | 0.02 | 0.24 | 0.08 | 0.02 – 0.09 | 0.09 – 0.39 | 3.11 | **0.002** |
| **Random Effects** | | | | | | | | |
| $\sigma^2$ | 0.02 | | | | | | | |
| $\tau_{00\ participant}$ | 0.02 | | | | | | | |
| ICC | 0.44 | | | | | | | |
| $N_{participant}$ | 92 | | | | | | | |
| Observations | 460 | | | | | | | |
| Marginal $R^2$ / Conditional $R^2$ | 0.206 / 0.552 | | | | | | | |

**Table S4. Linear model of effects of recognition-demand level and accuracy on confidence in young and older adults**

| | Confidence | | | | | | | | | |
|---|---|---|---|---|---|---|---|---|---|---|
| *Predictors* | *Estimates* | *std. Error* | *std. Beta* | *standardized std. Error* | *CI* | *standardized CI* | *Statistic* | *std. Statistic* | *p* | *std. p* |
| (Intercept) | 0.23 | 0.21 | 0.05 | 0.12 | -0.18 – 0.64 | -0.19 – 0.29 | 1.13 | 0.43 | 0.261 | 0.669 |
| Recognition-demand level | 0.07 | 0.02 | 0.02 | 0.04 | 0.03 – 0.11 | -0.07 – 0.10 | 3.71 | 0.38 | **<0.001** | 0.702 |
| Age group [Older adults] | 0.11 | 0.05 | 0.03 | 0.15 | 0.02 – 0.20 | -0.26 – 0.32 | 2.31 | 0.18 | **0.022** | 0.860 |
| Accuracy | 0.36 | 0.02 | 0.59 | 0.04 | 0.31 – 0.41 | 0.51 – 0.67 | 14.59 | 14.35 | **<0.001** | **<0.001** |
| Sex [Male] | -0.06 | 0.03 | -0.18 | 0.10 | -0.12 – 0.01 | -0.39 – 0.02 | -1.79 | -1.79 | 0.077 | 0.077 |
| Vocab score | 0.00 | 0.00 | 0.01 | 0.07 | -0.00 – 0.00 | -0.13 – 0.15 | 0.17 | 0.17 | 0.864 | 0.864 |
| Working memory score | 0.02 | 0.02 | 0.07 | 0.06 | -0.01 – 0.05 | -0.04 – 0.18 | 1.19 | 1.19 | 0.236 | 0.236 |
| Recognition-demand level × Age group [Older adults] | 0.00 | 0.02 | -0.10 | 0.05 | -0.05 – 0.05 | -0.20 – 0.00 | 0.07 | -1.90 | 0.941 | 0.058 |
| Recognition-demand level × Accuracy | -0.13 | 0.03 | -0.21 | 0.04 | -0.18 – -0.08 | -0.29 – -0.13 | -5.01 | -5.01 | **<0.001** | **<0.001** |
| Age group [Older adults] × Accuracy | -0.19 | 0.03 | -0.32 | 0.05 | -0.25 – -0.13 | -0.42 – -0.22 | -6.04 | -6.19 | **<0.001** | **<0.001** |
| Recognition-demand level × Age group [Older adults] × Accuracy | -0.06 | 0.03 | -0.10 | 0.05 | -0.12 – 0.00 | -0.20 – 0.00 | -1.91 | -1.91 | 0.056 | 0.056 |
| **Random Effects** | | | | | | | | | | |
| $\sigma^2$ | 0.05 | | | | | | | | | |
| $\tau_{00\ participant}$ | 0.02 | | | | | | | | | |

| | |
|---|---|
| ICC | 0.25 |
| N $_{participant}$ | 92 |
| Observations | 879 |
| Marginal $R^2$ / Conditional $R^2$ | 0.270 / 0.454 |

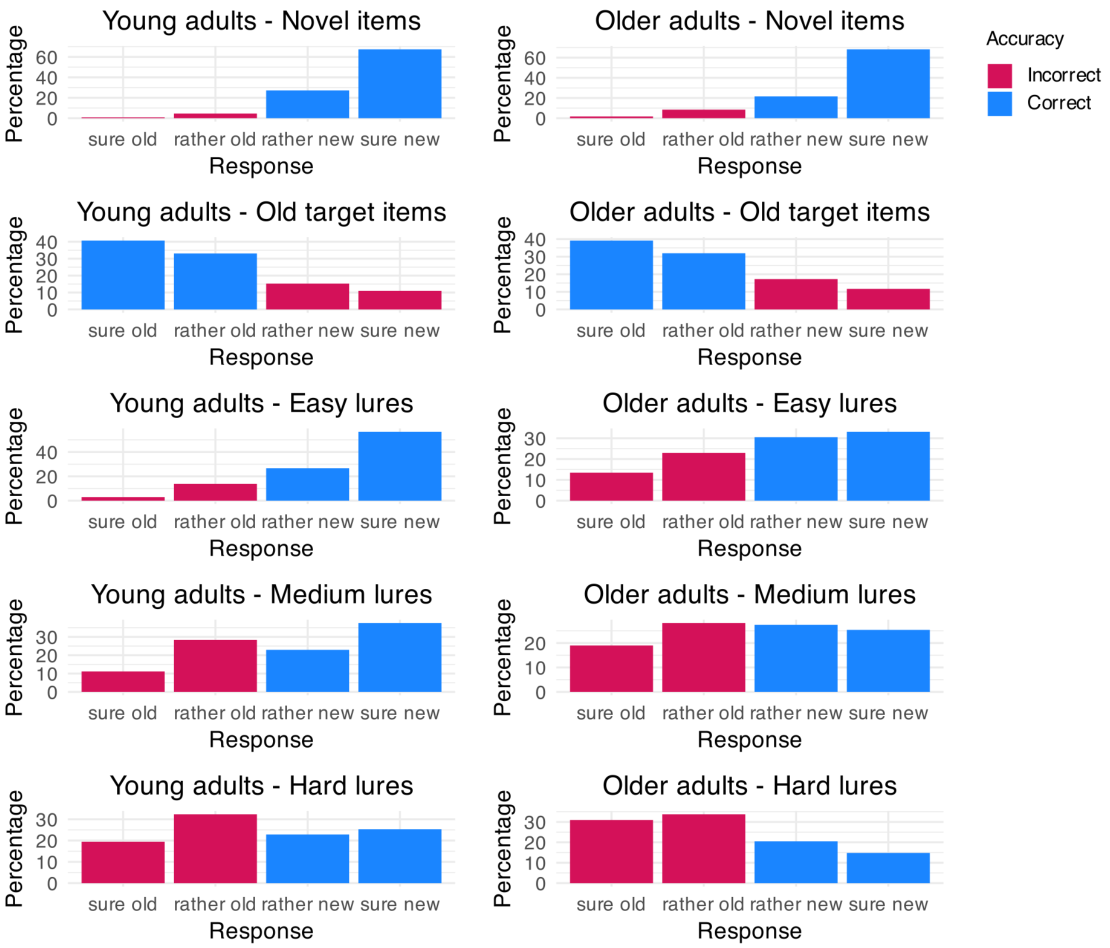


**Figure S1. Responses in confidence rating and accuracy thereof.** Responses given in the confidence rating during the recognition task by young and older adults for each recognition-demand level, including the distribution of correct and incorrect responses.

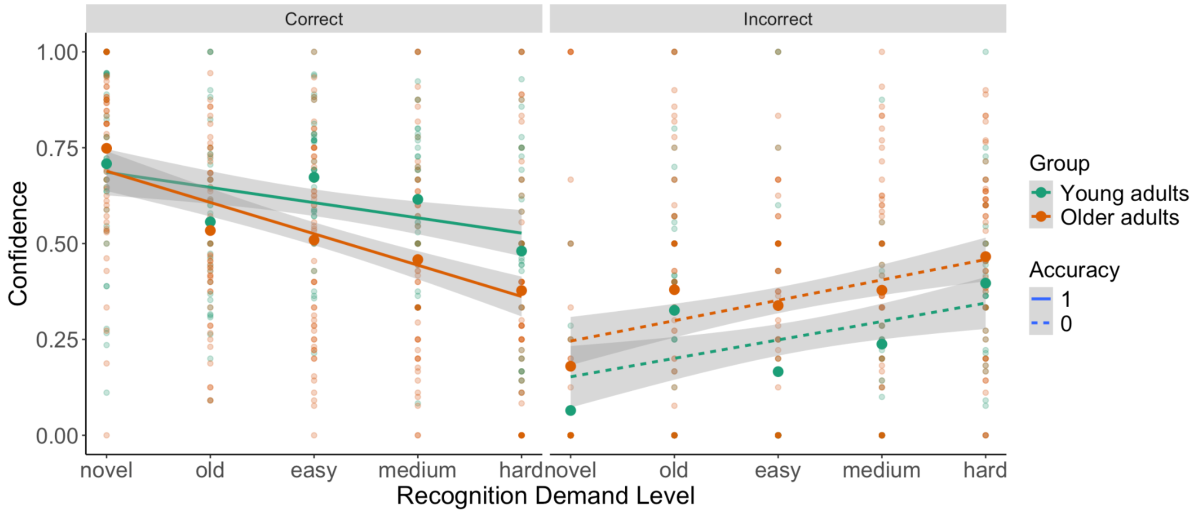


**Figure S2. Confidence by recognition-demand level, age group, and accuracy.**

**Table S5. Linear model of effects of recall-demand level on recall score in young and older adults**

| | Recall score | | | | | | | |
|---|---|---|---|---|---|---|---|---|
| *Predictors* | *Estimates* | *std. Error* | *std. Beta* | *standardized std. Error* | *CI* | *standardized CI* | *Statistic* | *p* |
| (Intercept) | 0.12 | 0.05 | 1.08 | 0.15 | 0.02 – 0.22 | 0.78 – 1.38 | 2.39 | **0.019** |
| Recall-demand level | -0.02 | 0.00 | -0.29 | 0.02 | -0.02 – -0.01 | -0.34 – -0.24 | -12.25 | **<0.001** |
| Age group [Older adults] | -0.08 | 0.01 | -1.46 | 0.19 | -0.11 – -0.06 | -1.82 – -1.09 | -7.86 | **<0.001** |
| Sex [Male] | -0.02 | 0.01 | -0.38 | 0.13 | -0.04 – -0.01 | -0.64 – -0.12 | -2.89 | **0.005** |
| Vocab score | -0.00 | 0.00 | -0.00 | 0.09 | -0.00 – 0.00 | -0.18 – 0.17 | -0.05 | 0.960 |
| Working memory score | 0.00 | 0.00 | 0.06 | 0.07 | -0.00 – 0.01 | -0.08 – 0.20 | 0.88 | 0.384 |
| Recall-demand level × Age group [Older adults] | 0.01 | 0.00 | 0.17 | 0.03 | 0.01 – 0.01 | 0.11 – 0.23 | 5.78 | **<0.001** |
| **Random Effects** | | | | | | | | |
| $\sigma^2$ | 0.00 | | | | | | | |
| $\tau_{00\ participant}$ | 0.00 | | | | | | | |

| | |
|---|---|
| ICC | 0.90 |
| $N_{participant}$ | 92 |
| Observations | 184 |
| Marginal $R^2$ / Conditional $R^2$ | 0.591 / 0.961 |

**Table S6. Linear model of effects of recall-demand level on recall error score in young and older adults**

| | Recall error score (log transformed) | | | | | | | | | | | |
|---|---|---|---|---|---|---|---|---|---|---|---|---|
| *Predictors* | *Estimates* | *std. Error* | *std. Beta* | *exp Beta* | *% change* | *standardized std. Error* | *CI* | *standardized CI* | *Statistic* | *std. Statistic* | *p* | *std. p* |
| (Intercept) | -2.09 | 1.12 | 0.26 | 0.124 | -87.60 | 0.09 | -4.31 – 0.14 | 0.09 – 0.43 | -1.86 | 3.03 | 0.066 | **0.003** |
| Age group [Older adults] | 0.93 | 0.24 | 0.34 | 2.52 | 152.00 | 0.11 | 0.46 – 1.40 | 0.13 – 0.55 | 3.92 | 3.22 | **<0.001** | **0.002** |
| Sex [Male] | 0.19 | 0.17 | 0.11 | 1.21 | 20.50 | 0.08 | -0.15 – 0.52 | -0.04 – 0.26 | 1.10 | 1.45 | 0.273 | 0.150 |
| Vocab score | 0.01 | 0.01 | -0.00 | 1.01 | 0.54 | 0.05 | -0.02 – 0.03 | -0.10 – 0.10 | 0.47 | -0.05 | 0.639 | 0.963 |
| Working memory score | -0.33 | 0.10 | -0.14 | 0.72 | -27.80 | 0.04 | -0.52 – -0.13 | -0.22 – -0.06 | -3.36 | -3.38 | **0.001** | **0.001** |
| Observations | 89 | | | | | | | | | | | |
| $R^2$ / $R^2$ adjusted | 0.394 / 0.365 | | | | | | | | | | | |

Three older individuals were excluded for this analysis, as there were methodological problems with their recall error scoring.

**Table S7. Linear model of effects of recognition-demand level on accuracy in older adults**

| | Accuracy |
|---|---|

| *Predictors* | *Estimates* | *std. Error* | *std. Beta* | *standardized std. Error* | *CI* | *standardized CI* | *Statistic* | *p* |
|---|---|---|---|---|---|---|---|---|
| (Intercept) | 0.57 | 0.13 | 0.07 | 0.06 | 0.32 – 0.82 | -0.04 – 0.19 | 4.51 | **<0.001** |
| Recognition-demand level | -0.17 | 0.01 | -0.70 | 0.04 | -0.19 – -0.15 | -0.78 – -0.61 | -15.65 | **<0.001** |
| *APOE4* Group [Carrier] | -0.02 | 0.02 | -0.09 | 0.10 | -0.07 – 0.02 | -0.28 – 0.10 | -0.92 | 0.360 |
| Alzheimer's pathology | 0.01 | 0.01 | 0.05 | 0.04 | -0.01 – 0.03 | -0.03 – 0.14 | 1.16 | 0.245 |
| Age at baseline | 0.05 | 0.03 | 0.06 | 0.04 | -0.01 – 0.12 | -0.01 – 0.14 | 1.65 | 0.099 |
| Sex [Male] | -0.03 | 0.02 | -0.11 | 0.08 | -0.06 – 0.01 | -0.26 – 0.05 | -1.37 | 0.172 |
| Vocab score | 0.00 | 0.00 | 0.01 | 0.04 | -0.00 – 0.00 | -0.07 – 0.10 | 0.31 | 0.756 |
| Working memory score | 0.03 | 0.01 | 0.15 | 0.04 | 0.01 – 0.05 | 0.06 – 0.23 | 3.48 | **0.001** |
| Recognition-demand level × *APOE4* Group [Carrier] | -0.04 | 0.02 | -0.18 | 0.09 | -0.09 – -0.00 | -0.36 – -0.01 | -2.06 | **0.041** |
| **Random Effects** | | | | | | | | |
| $\sigma^2$ | 0.02 | | | | | | | |
| $\tau_{00\ participant}$ | 0.00 | | | | | | | |
| $N_{participant}$ | 56 | | | | | | | |
| Observations | 280 | | | | | | | |
| Marginal $R^2$ / Conditional $R^2$ | 0.586 / NA | | | | | | | |

Exploratory post-hoc analysis of the interaction effect with recognition-demand level as a factor revealed a significant difference between age groups for the fifth recognition-demand level (hard lures) (t(295) = 2.465, $p$ = 0.0143, SE = 0.049) using emmeans.

**Table S8. Linear model of effects of recognition-demand level on confidence in older adults**

| | Confidence | | | | | | | |
|---|---|---|---|---|---|---|---|---|
| *Predictors* | *Estimates* | *std. Error* | *std. Beta* | *standardized std. Error* | *CI* | *standardized CI* | *Statistic* | *p* |
| (Intercept) | 0.26 | 0.27 | -0.06 | 0.14 | -0.27 – 0.80 | -0.33 – 0.22 | 0.99 | 0.324 |
| Recognition-demand level | -0.08 | 0.01 | -0.36 | 0.04 | -0.10 – -0.06 | -0.45 – -0.28 | -8.25 | **<0.001** |
| *APOE4* Group [Carrier] | 0.00 | 0.05 | 0.01 | 0.24 | -0.10 – 0.11 | -0.46 – 0.49 | 0.05 | 0.957 |
| Recognition-demand level U-shape | 0.05 | 0.01 | 0.25 | 0.05 | 0.03 – 0.08 | 0.14 – 0.35 | 4.65 | **<0.001** |
| Alzheimer's pathology | -0.01 | 0.02 | -0.04 | 0.10 | -0.05 – 0.03 | -0.23 – 0.15 | -0.43 | 0.671 |
| Age at baseline | -0.02 | 0.06 | -0.03 | 0.09 | -0.15 – 0.11 | -0.20 – 0.15 | -0.31 | 0.761 |
| Sex [Male] | -0.11 | 0.04 | -0.47 | 0.17 | -0.18 – -0.03 | -0.82 – -0.13 | -2.74 | **0.008** |
| Vocab score | 0.00 | 0.00 | 0.10 | 0.10 | -0.00 – 0.01 | -0.09 – 0.29 | 1.06 | 0.296 |
| Working memory score | 0.02 | 0.02 | 0.10 | 0.09 | -0.02 – 0.06 | -0.09 – 0.29 | 1.06 | 0.296 |
| Recognition-demand level × *APOE4* Group [Carrier] | 0.01 | 0.02 | 0.05 | 0.09 | -0.03 – 0.05 | -0.12 – 0.23 | 0.58 | 0.560 |
| Recognition-demand level U-shape × *APOE4* Group [Carrier] | 0.02 | 0.02 | 0.11 | 0.11 | -0.02 – 0.07 | -0.10 – 0.32 | 1.04 | 0.298 |
| **Random Effects** | | | | | | | | |
| $\sigma^2$ | 0.02 | | | | | | | |
| $\tau_{00\ participant}$ | 0.02 | | | | | | | |
| ICC | 0.44 | | | | | | | |
| $N_{participant}$ | 56 | | | | | | | |
| Observations | 280 | | | | | | | |
| Marginal $R^2$ / Conditional $R^2$ | 0.268 / 0.591 | | | | | | | |

**Table S9. Linear model of effects of recognition-demand level and accuracy on confidence in older adults**

| | Confidence | | | | | | | | | |
|---|---|---|---|---|---|---|---|---|---|---|
| *Predictors* | *Estimates* | *std. Error* | *std. Beta* | *standardized std. Error* | *CI* | *standardized CI* | *Statistic* | *std. Statistic* | *p* | *std. p* |
| (Intercept) | 0.28 | 0.27 | 0.17 | 0.10 | -0.26 – 0.81 | -0.04 – 0.37 | 1.03 | 1.64 | 0.307 | 0.107 |
| Recognition-demand level | 0.05 | 0.02 | -0.10 | 0.04 | 0.01 – 0.08 | -0.18 – -0.03 | 2.79 | -2.71 | **0.005** | **0.007** |
| *APOE4* Group [Carrier] | -0.00 | 0.05 | -0.08 | 0.17 | -0.11 – 0.11 | -0.41 – 0.26 | -0.06 | -0.45 | 0.949 | 0.657 |
| Accuracy | 0.18 | 0.02 | 0.29 | 0.04 | 0.13 – 0.22 | 0.22 – 0.36 | 8.00 | 7.70 | **<0.001** | **<0.001** |
| Alzheimer’s pathology | -0.00 | 0.02 | -0.01 | 0.08 | -0.05 – 0.04 | -0.16 – 0.15 | -0.07 | -0.07 | 0.944 | 0.944 |
| Age at baseline | -0.01 | 0.07 | -0.01 | 0.07 | -0.14 – 0.13 | -0.14 – 0.13 | -0.10 | -0.10 | 0.919 | 0.919 |
| Sex [Male] | -0.10 | 0.04 | -0.35 | 0.14 | -0.18 – -0.02 | -0.62 – -0.07 | -2.55 | -2.55 | **0.014** | **0.014** |
| Vocab score | 0.00 | 0.00 | 0.04 | 0.07 | -0.00 – 0.01 | -0.11 – 0.19 | 0.52 | 0.52 | 0.605 | 0.605 |
| Working memory score | 0.01 | 0.02 | 0.05 | 0.07 | -0.03 – 0.05 | -0.10 – 0.20 | 0.65 | 0.65 | 0.520 | 0.520 |
| Recognition-demand level × *APOE4* Group [Carrier] | 0.11 | 0.03 | 0.07 | 0.07 | 0.04 – 0.17 | -0.07 – 0.22 | 3.29 | 0.97 | **0.001** | 0.335 |
| Recognition-demand level × Accuracy | -0.15 | 0.02 | -0.25 | 0.04 | -0.19 – -0.11 | -0.32 – -0.17 | -6.58 | -6.58 | **<0.001** | **<0.001** |
| *APOE4* Group [Carrier] × Accuracy | -0.04 | 0.04 | -0.08 | 0.07 | -0.13 – 0.05 | -0.22 – 0.07 | -0.86 | -1.04 | 0.391 | 0.301 |

| | | | | | | | | | |
|---|---|---|---|---|---|---|---|---|---|
| Recognition-demand level × *APOE4* Group [Carrier] × Accuracy | -0.17 | 0.05 | -0.28 | 0.07 | -0.25 – -0.08 | -0.42 – -0.13 | -3.70 | -3.70 | **<0.001** | **<0.001** |

**Random Effects**

| | |
|---|---|
| $\sigma^2$ | 0.05 |
| $\tau_{00\ participant}$ | 0.02 |
| ICC | 0.25 |
| $N_{participant}$ | 56 |
| Observations | 541 |
| Marginal $R^2$ / Conditional $R^2$ | 0.244 / 0.431 |

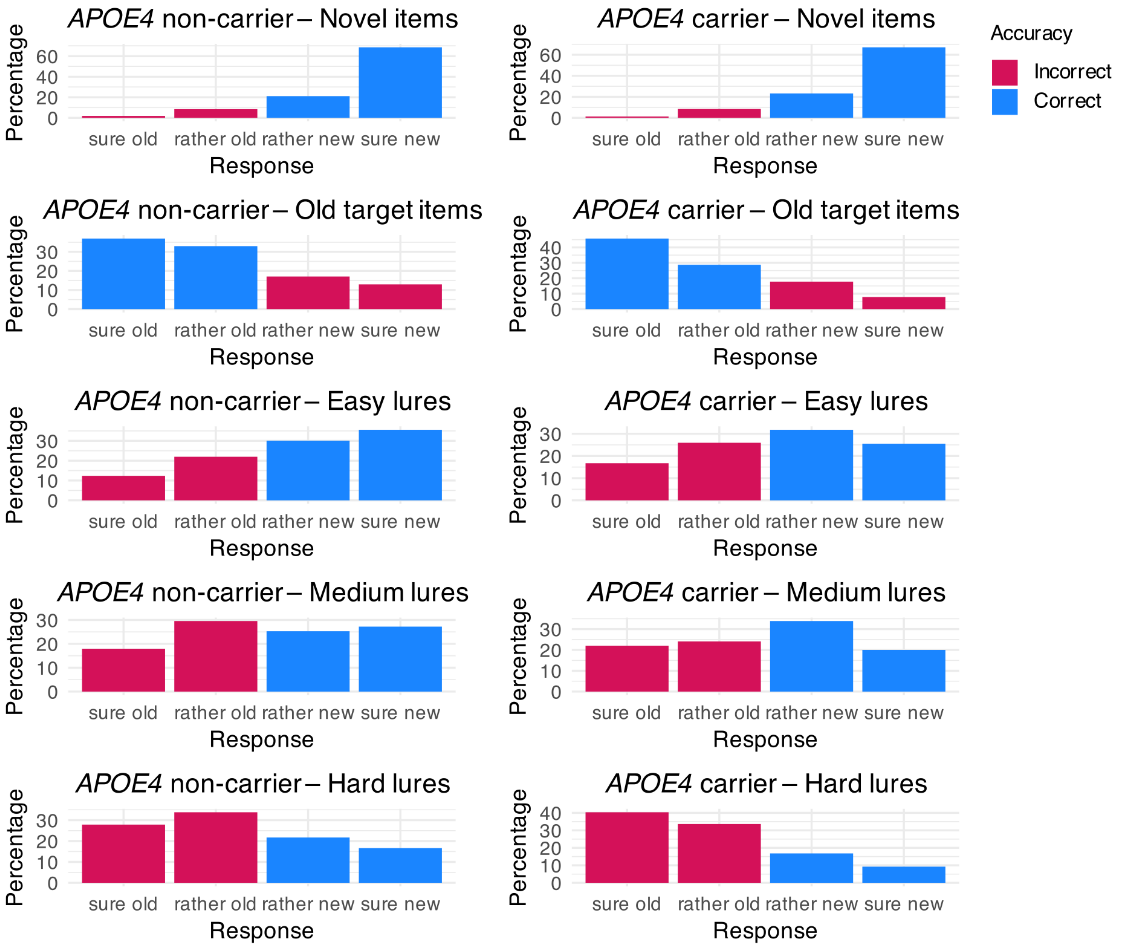


**Figure S3. Responses in confidence rating and accuracy thereof.** Responses given in the confidence rating during the recognition task by older *APOE4* carriers and non-carriers for each recognition-demand level, including the distribution of correct and incorrect responses.

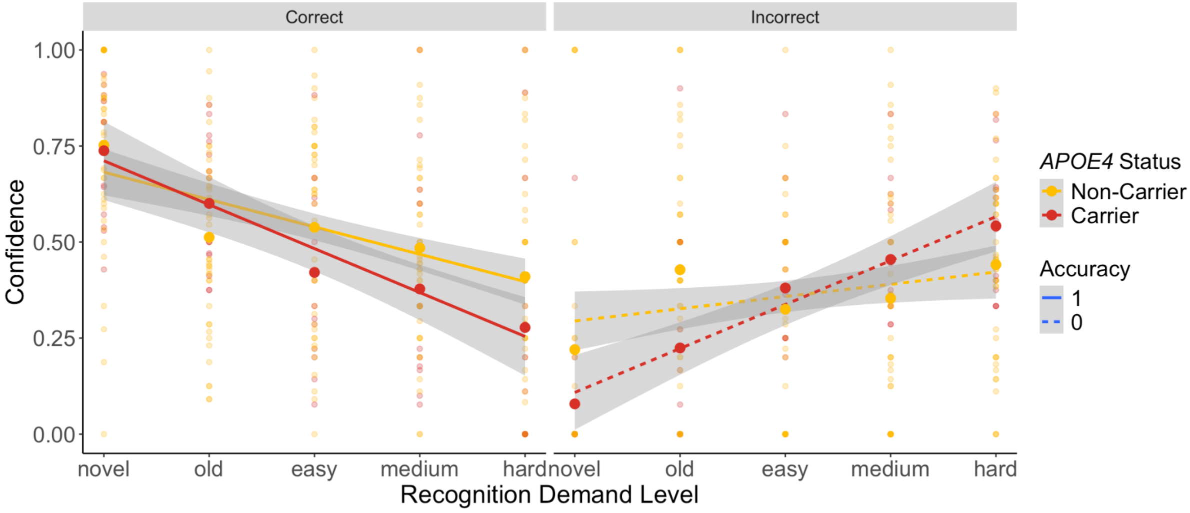


**Figure S4. Confidence by recognition-demand level, *APOE4* group, and accuracy.**

### Table S10. Linear model of effects on VLMT recognition in older adults

| | VLMT recognition accuracy | | | | | | | |
|---|---|---|---|---|---|---|---|---|
| *Predictors* | *Estimates* | *std. Error* | *std. Beta* | *standardized std. Error* | *CI* | *standardized CI* | *Statistic* | *p* |
| (Intercept) | 0.96 | 0.17 | 0.26 | 0.21 | 0.62 – 1.31 | -0.15 – 0.68 | 5.60 | **<0.001** |
| *APOE4* Group [Carrier] | -0.03 | 0.03 | -0.37 | 0.34 | -0.10 – 0.03 | -1.06 – 0.33 | -1.06 | 0.294 |
| Alzheimer's pathology | -0.00 | 0.01 | -0.03 | 0.15 | -0.03 – 0.03 | -0.34 – 0.28 | -0.20 | 0.842 |
| Age at baseline | -0.02 | 0.04 | -0.08 | 0.14 | -0.11 – 0.06 | -0.36 – 0.20 | -0.57 | 0.572 |
| Sex [Male] | -0.03 | 0.03 | -0.37 | 0.28 | -0.09 – 0.02 | -0.93 – 0.19 | -1.34 | 0.186 |
| Vocab score | -0.00 | 0.00 | -0.01 | 0.15 | -0.00 – 0.00 | -0.31 – 0.30 | -0.03 | 0.973 |
| Working memory score | 0.00 | 0.01 | 0.03 | 0.15 | -0.02 – 0.03 | -0.28 – 0.33 | 0.17 | 0.862 |
| Observations | 56 | | | | | | | |
| $R^2$ / $R^2$ adjusted | 0.083 / -0.029 | | | | | | | |

**Table S11. Linear model of effects on RCFT recognition in older adults**

| | RCFT recognition accuracy | | | | | | | |
|---|---|---|---|---|---|---|---|---|
| *Predictors* | *Estimates* | *std. Error* | *std. Beta* | *standardized std. Error* | *CI* | *standardized CI* | *Statistic* | *p* |
| (Intercept) | 1.00 | 0.11 | -0.08 | 0.19 | 0.78 – 1.22 | -0.47 – 0.30 | 9.20 | **<0.001** |
| *APOE4* Group [Carrier] | 0.01 | 0.02 | 0.18 | 0.32 | -0.03 – 0.05 | -0.46 – 0.83 | 0.57 | 0.572 |
| Alzheimer's pathology | -0.03 | 0.01 | -0.47 | 0.14 | -0.05 – -0.01 | -0.76 – -0.18 | -3.30 | **0.002** |
| Age at baseline | 0.02 | 0.03 | 0.11 | 0.13 | -0.03 – 0.08 | -0.15 – 0.37 | 0.87 | 0.389 |
| Sex [Male] | 0.00 | 0.02 | 0.08 | 0.26 | -0.03 – 0.04 | -0.44 – 0.60 | 0.30 | 0.764 |
| Vocab score | -0.00 | 0.00 | -0.22 | 0.14 | -0.00 – 0.00 | -0.51 – 0.07 | -1.55 | 0.128 |
| Working memory score | 0.01 | 0.01 | 0.10 | 0.14 | -0.01 – 0.02 | -0.18 – 0.39 | 0.74 | 0.462 |
| Observations | 56 | | | | | | | |
| $R^2$ / $R^2$ adjusted | 0.208 / 0.111 | | | | | | | |

**Table S12. Linear model of effects of recall-demand level on recall score in older adults**

| | Recall score | | | | | | | |
|---|---|---|---|---|---|---|---|---|
| *Predictors* | *Estimates* | *std. Error* | *std. Beta* | *standardized std. Error* | *CI* | *standardized CI* | *Statistic* | *p* |
| (Intercept) | 0.03 | 0.03 | 0.29 | 0.15 | -0.03 – 0.10 | -0.01 – 0.59 | 1.00 | 0.320 |
| Recall-demand level | -0.01 | 0.00 | -0.29 | 0.04 | -0.01 – -0.00 | -0.38 – -0.21 | -6.73 | **<0.001** |
| *APOE4* Group [Carrier] | 0.00 | 0.01 | 0.18 | 0.25 | -0.01 – 0.02 | -0.32 – 0.68 | 0.71 | 0.483 |
| Alzheimer's pathology | -0.00 | 0.00 | -0.12 | 0.11 | -0.01 – 0.00 | -0.34 – 0.10 | -1.08 | 0.284 |

| | | | | | | | | |
|---|---|---|---|---|---|---|---|---|
| Age at baseline | -0.02 | 0.01 | -0.20 | 0.10 | -0.03 – 0.00 | -0.40 – 0.01 | -1.95 | 0.056 |
| Sex [Male] | -0.02 | 0.00 | -0.72 | 0.20 | -0.03 – -0.01 | -1.12 – -0.32 | -3.57 | **0.001** |
| Vocab score | 0.00 | 0.00 | 0.04 | 0.11 | -0.00 – 0.00 | -0.18 – 0.26 | 0.33 | 0.739 |
| Working memory score | 0.01 | 0.00 | 0.24 | 0.11 | 0.00 – 0.01 | 0.02 – 0.46 | 2.21 | **0.031** |
| Recall-demand level × *APOE4* Group [Carrier] | 0.00 | 0.00 | 0.03 | 0.09 | -0.00 – 0.00 | -0.15 – 0.20 | 0.33 | 0.745 |
| **Random Effects** | | | | | | | | |
| $\sigma^2$ | 0.00 | | | | | | | |
| $\tau_{00\ participant}$ | 0.00 | | | | | | | |
| ICC | 0.74 | | | | | | | |
| $N_{participant}$ | 56 | | | | | | | |
| Observations | 112 | | | | | | | |
| Marginal $R^2$ / Conditional $R^2$ | 0.376 / 0.841 | | | | | | | |

### Table S13. Linear model of effects of recall-demand level on recall error score in older adults

| | Recall error score (log transformed) | | | | | | | | | | | |
|---|---|---|---|---|---|---|---|---|---|---|---|---|
| *Predictors* | *Estimates* | *std. Error* | *std. Beta* | *exp Beta* | *% change* | *standardized std. Error* | *CI* | *standardized CI* | *Statistic* | *std. Statistic* | *p* | *std. p* |
| (Intercept) | -0.52 | 1.45 | 0.54 | 0.596 | -40.40 | 0.09 | -3.43 – 2.39 | 0.37 – 0.71 | -0.36 | 6.30 | 0.722 | **<0.001** |
| *APOE4* Group [Carrier] | -0.08 | 0.27 | -0.06 | 0.925 | -7.55 | 0.15 | -0.62 – 0.46 | -0.36 – 0.24 | -0.29 | -0.39 | **0.772** | **0.699** |
| Alzheimer's pathology | -0.03 | 0.12 | -0.02 | 0.967 | -3.33 | 0.07 | -0.27 – 0.20 | -0.16 – 0.11 | -0.29 | -0.38 | 0.770 | 0.706 |
| Age at baseline | 0.47 | 0.37 | 0.08 | 1.60 | 60.30 | 0.06 | -0.26 – 1.21 | -0.04 – 0.20 | 1.29 | 1.26 | 0.203 | 0.213 |

| | | | | | | | | | | | | |
|---|---|---|---|---|---|---|---|---|---|---|---|---|
| Sex [Male] | 0.12 | 0.22 | 0.09 | 1.13 | 12.50 | 0.12 | -0.32 – 0.55 | -0.15 – 0.33 | 0.54 | 0.77 | **0.588** | **0.447** |
| Vocab score | -0.00 | 0.01 | -0.02 | 0.998 | -0.24 | 0.12 | -0.32 – 0.55 | -0.15 – 0.33 | 0.54 | 0.77 | **0.588** | **0.447** |
| Working memory score | -0.31 | 0.12 | -0.17 | 0.736 | -26.40 | 0.06 | -0.03 – 0.02 | -0.15 – 0.11 | -0.19 | -0.31 | **0.849** | **0.754** |
| Observations | 53 | | | | | | | | | | | |
| $R^2$ / $R^2$ adjusted | 0.199 / 0.095 | | | | | | | | | | | |

Three older individuals were excluded from this analysis, as there were methodological problems with their recall error scoring.

### References of supplementary


Bird, C. M., Keidel, J. L., Ing, L. P., Horner, A. J., & Burgess, N. (2015). Consolidation of Complex Events via Reinstatement in Posterior Cingulate Cortex. *Journal of Neuroscience*, *35*(43), 14426–14434.

Burke, A., Heuer, F., & Reisberg, D. (1992). Remembering emotional events. *Memory & Cognition*, *20*(3). https://doi.org/10.3758/bf03199665

Wessel, I., Zandstra, A. R. E., Hengeveld, H. M. E., & Moulds, M. L. (2015). Collaborative recall of details of an emotional film. *Memory (Hove, England)*, *23*(3), 437–444.
